# Cardiovascular exercise mitigates inflammation in the early subacute phase of stroke recovery: a secondary analysis of a randomized controlled trial

**DOI:** 10.64898/2026.09.28.26364152

**Authors:** Bernat De Las Heras, Lynden Rodrigues, Kevin Moncion, Anke Van Roy, Madhura Lotlikar, Fotini Michalakis, Kira Sikorska, Adam Sutoski, Ashanté Bon, Nathalie Arbour, Mark Bayley, Janice Eng, Marc Roig

**Author notes:** **Corresponding author:** Marc Roig, Memory and Motor Rehabilitation Laboratory (MEMORY-LAB), Jewish Rehabilitation Hospital, 3205 Place Alton-Goldbloom, Laval, QC H7V 1R2, Canada.

## Abstract

**Background:** Excessive inflammation in early phases post-ischemic stroke is associated with poor long-term outcome and increased stroke recurrence. Interventions capable of downregulating persistent states of inflammation could improve recovery and prevent secondary stroke. Cardiovascular exercise (CE) has well-established anti-inflammatory effects but its capacity to attenuate the exaggerated inflammatory response triggered by ischemic stroke is unclear.

**Methods:** We investigated the impact of eight weeks of CE on the serum concentration of inflammatory mediators in the early subacute phase post-stroke. Seventy-one patients with first-ever ischemic stroke were randomly assigned to moderate-to-vigorous CE (n=45) or standard care (SC) (n=26). Concentrations of CRP, IL-6, IL-8, IL-10, TNF-α and IL-1β were measured at baseline and after the interventions. Differences between groups were analyzed with linear mixed models adjusted for age and sex.

**Results:** Compared with the CE group, the SC group showed significant increases in IL-6 (β=0.585, 95% CI=0.164–1.006, p=0.0073), IL-8 (β=0.160, 95% CI=0.004–0.317, p=0.0447), and TNF-α (β=0.0845, 95% CI=0.0137–0.1553, p=0.0206). The SC group also showed greater increases in CRP (β=0.250, 95% CI=−0.407, 0.906, p=0.4499) and IL-10 (β=0.251, 95% CI=−0.024, 0.527, p=0.0729) but group differences were not significant. IL-1β levels remained stable and differences between groups were not significant (β=−0.008, 95% CI=−0.058, 0.042, p=0.7618).

**Conclusions:** When introduced in the early subacute phase of stroke recovery, moderate-to-vigorous CE mitigates increases in inflammation, but the effect differs across mediators. To what extent reducing the inflammatory response in the early stages post-stroke with CE can impact long-term recovery and stroke recurrence needs to be determined.

## 1. Introduction

Brain inflammation in response to ischemia regulates the immune and neuroplastic response after stroke (Kriz and Lalancette-Hébert, 2009; Shi et al., 2019). Pro-inflammatory mediators such as c-reactive protein (CRP), interleukins (IL) 6, 8, 1β and tumor necrosis factor alpha (TNF-α), as well as anti-inflammatory mediators such as IL-10, have been shown to play an active role as neuromodulators by engaging bidirectionally with the central nervous system and participating in reparative processes following brain injury (Iordache et al., 2025). The inflammatory and immune response during the acute phase post-stroke (1-7 days) is needed for neovascularization, tissue repair and neuroplasticity (Gertz et al., 2012). However, the prolonged overexpression of pro-inflammatory mediators in later phases post-stroke can result in detrimental effects for long-term recovery via alterations in neural function and neuroplasticity (Zhu et al., 2022). Importantly, persistent excessive inflammation in early phases of stroke recovery is also associated with an increased risk of suffering a secondary stroke (Kelly et al., 2021).

Several studies have investigated how the concentration of inflammatory mediators in the early stages post-stroke correlate with long-term disability outcomes (Couch et al., 2022; Lai et al., 2019). There is evidence that abnormally elevated CRP levels measured the days following a stroke are associated with reduced long-term recovery (Geng et al., 2016; Winbeck et al., 2002) and increased stroke recurrence (Zietz et al., 2024). Increased IL-6 and TNF-α levels have also been linked to poorer functional outcomes both at discharge (Băcilă et al., 2025) and three months post-stroke (Aref et al., 2020). Higher IL-8 levels are associated with disability post-stroke (Shaheen et al., 2018) and blocking IL-1β action may enhance recovery (Grayston et al., 2025; Sobowale et al., 2016). Conversely, although more evidence is needed, lower levels of the anti-inflammatory cytokine IL-10 measured 24 hours after admission have been linked with poor functional recovery (Sun et al., 2021). Importantly, associations between the concentration of these inflammatory mediators and recovery and recurrence outcomes have also been observed with blood samples taken several months after stroke (McCabe et al., 2024; Sandvig et al., 2023), suggesting that modulating inflammation in later phases of stroke recovery might be also important. Since the inflammatory response triggered by the ischemic stroke is proportional to the amount of neural damage produced (e.g., infarct size) (Pacinella et al., 2025), one could argue that inflammation is simply an epiphenomenon, whose association with disability is explained by the fact that more inflammation simply reflects more tissue damage and thus poorer long-term recovery. However, recent studies have demonstrated that pharmacologically modulating the activity of some of these pro-inflammatory mediators (e.g., reducing IL-6 signaling activity) could have positive therapeutic effects (Iordache et al., 2025). Hence, while the underlying mechanisms are certainly complex and some aspects remain unknown, inflammation plays an active role in regulating post-stroke recovery and interventions modulating it could improve patients’ outcomes (Clausen et al., 2020; Di Filippo et al., 2008; Zietz et al., 2024).

One easy to implement and affordable intervention that has been shown to have significant anti-inflammatory effects in individuals without neurological injury is cardiovascular exercise (CE) (Poorhabibi et al., 2025; Tayebi et al., 2025). Indeed, the positive effect of physical activity in general (Beavers et al., 2010), and CE in particular, on mitigating systemic inflammation via reductions of pro-inflammatory (CRP, IL-6, IL-8, TNF-α, IL-1β) and increases in anti-inflammatory (IL-10) mediators in non-clinical populations has been well established (Tayebi et al., 2025). However, there is still sparse data regarding the capacity of CE to reduce the exacerbated inflammatory response usually observed post-ischemic stroke (Bitencourt et al., 2025), especially in the early phases of subacute recovery (7 days to 3 months), when modulating inflammation to ensure an appropriate neuroplastic response could have substantial therapeutic effects (Murphy and Corbett, 2009) and, potentially, reduce the risk of stroke recurrence (Zietz et al., 2024).

After the acute phase post-stroke (1-7 days post-stroke), during which the inflammatory and immune response is particularly prominent (Iordache et al., 2025), the early (7 days to 3 months) and late (3 to 6 months) subacute phases of recovery follow (Bernhardt et al., 2017). Given the heightened window of neuroplasticity opened in the early subacute phase (Biernaskie et al., 2004) and the fact that the risk of stroke recurrence remains elevated the first three months after stroke (Stahmeyer et al., 2019), reducing inflammation in this phase of stroke recovery with CE could be particularly important to optimize rehabilitation outcomes (Dromerick et al., 2021) and improve long-term secondary prevention (Zietz et al., 2024).

The goal of this randomized controlled trial (RCT) was to examine the effect of eight weeks of progressive moderate-to-vigorous CE plus standard care (SC), compared to SC alone, on biomarkers of neuroplasticity (De Las Heras et al., 2024) such as brain derived neurotrophic factor (BDNF) (De Las Heras et al., 2025) as well as the modulatory effect of the Val66Met polymorphism on the serum BDNF response to CE (De Las Heras et al., 2022) in individuals at the early subacute phase of recovery. Given the deleterious effect of inflammation on BDNF expression and secretion (Calabrese et al., 2014), and the paucity of studies investigating the effects of CE on inflammation in early stages of subacute recovery (Bitencourt et al., 2025), we performed a secondary analysis of the RCT that investigated changes in pro-(CRP, IL-6, IL-8, TNF-α, IL-1β) and anti-inflammatory mediators (IL-10) in response to CE. We hypothesized that compared with SC alone, CE plus SC would attenuate systemic inflammation, reducing the concentration of CRP, IL-6, IL-8, TNF-α and IL-1β and increasing the concentration of IL-10.

## 2. Methods

### 2.1 Design

The secondary analysis presented in this manuscript, which was not pre-specified in the original study registration (ClinicalTrials.gov: NCT05076747), use blood samples collected in a two-arm parallel group RCT (De Las Heras et al., 2025). Following baseline assessments, participants were randomized (2:1, variable block sizes) to either an eight-week CE plus SC or SC alone. The reason for this unbalanced randomization was the unequal allelic frequency of the Val66Met polymorphism across populations (Petryshen et al., 2010). To ensure adequate power to detect genotype-related differences in BDNF response in the exercise group, participants were randomized using a 2:1 ratio (De Las Heras et al., 2025).

The block allocation sequence was generated and concealed by the principal investigator (MR). Outcome data were collected at baseline (T0) and after the intervention period (T1). Blood analyses were performed by technicians blinded to the allocation of participants to the two arms of the study. Ethics approval was obtained from the local ERB (CRIR-1265-0817) and all participants provided written informed consent. Additional methodological details of this study, including the sample size estimation for the main outcome, can be found in the registered RCT and in the manuscript reporting one of the main outcomes (De Las Heras et al., 2025). This RCT adheres to the Consolidated Standards of Reporting Trials (CONSORT) guidelines. All data used in the analyses are available upon request.

### 2.2 Participants

Eligible participants were 40–80 years old and, at the time of enrollment in the study, within three months following first-ever ischemic stroke as confirmed by MRI/CT. Participants recruited were either hospital in-patients or early out-patients, and some patients transitioned from in-patient to out-patient rehabilitation services during participation in the study. They needed to be free of upper-limb musculoskeletal or neurological conditions other than stroke and have the capacity to follow instructions and perform the CE intervention safely. Participants were excluded if they had hemorrhagic stroke, cognitive impairment affecting informed consent, absolute contraindications to exercise, or were concurrently enrolled in another structured exercise training program outside the study.

### 2.3 Interventions

#### 2.3.1 Cardiovascular exercise (CE)

Besides receiving SC (see next section), participants in the CE group exercised three days/week × eight weeks. Training was conducted at the Jewish Rehabilitation Hospital (Quebec, Canada) on a whole-body recumbent stepper and delivered by a kinesiologist to one patient at a time. Weeks one to four involved a moderate-to-vigorous intensity continuous training protocol that targeted 65% to 80% of peak power output (PPO) for 20 to 35 minutes. Weeks five to eight followed a high-intensity interval training program, that involved eight × one-minute intervals of high-intensity exercise, interspersed with seven × one-minute low-intensity intervals. The high-intensity intervals targeted 85% PPO and were increased by 5% each week until reaching 100% PPO. Low intensity intervals targeted 35% PPO. Each session included 2.5-minute warm-up and cool down periods targeting 35% of PPO. Training prescriptions were individualized using a graded exercise test (GXT) (Moncion et al., 2025) performed at baseline, and at the end of week four to update training targets.

#### 2.3.2 Standard care (SC)

Standard care (SC) consisted of rehabilitation sessions conducted within the same rehabilitation center as the CE intervention (Jewish Rehabilitation Hospital) and it was prescribed by the stroke clinical unit. In addition to routine health monitoring by physicians and nursing staff, SC included physiotherapy, occupational therapy, and speech therapy sessions. The content and amount of rehabilitation, which varied among patients, was tailored to individual needs as determined by the stroke clinicians, with each therapy session lasting ∼45 minutes. To quantify potential divergences between groups in SC, we recorded the type and number of therapy sessions received by each patient from the beginning of the study to its conclusion.

### 2.4 Assessments

#### 2.4.1 General assessments

Participants’ age, biological sex, body mass index (BMI), medical history, including comorbidities (age-adjusted Charlson Comorbidity Index -CCI-) (Charlson ME, 1987) and stroke characteristics were collected at T0. Cognition (i.e., Montreal Cognitive Assessment, MoCA) (Dong et al., 2010), degree of neurological (i.e., National Institute of Health Stroke Severity, NIHSS) (Kwah and Diong, 2014) and motor deficit (i.e., Fugl-Meyer) (Lin JH, 2004), and cardiorespiratory fitness (Moncion et al., 2025) were collected at T0 and T1. Self-reported physical activity levels were measured at each time point using the physical activity scale for people with disabilities (PASIPD) and converted to metabolic equivalents (METs hour/day) (Washburn et al., 2002). Any adverse event, defined as any undesired experience that may present during the interventions, was registered during the duration of the study. Participants were asked not to change their physical activity habits outside the study and not to engage in moderate- or high-intensity physical activity 24 hours before any of the assessments.

#### 2.4.2 Cardio-respiratory fitness

Measurement of peak oxygen uptake (VO_2_peak in mL.Kg^-1^.min^-1^) with a GXT is the gold standard method for determining cardiorespiratory fitness (ACSM, 2024). A symptom-limited GXT utilizing a protocol validated for individuals across the stroke recovery continuum was performed on a whole-body recumbent stepper (NuStep T4r, Michigan, USA) (Moncion et al., 2025). During the GXT, heart rate (HR) was measured continuously while blood pressure (BP) and rate of perceived exertion (RPE) were recorded every two minutes. The GXT was also used to determine maximal HR (HR_max_) and PPO expressed in Watts. Indications for test termination followed current guidelines (ACSM, 2024).

#### 2.4.3 Blood collection and analysis

Blood collection was carried out by a hospital nurse. An antecubital intravenous line was placed in the non-paretic arm, with a waste sample collected before each blood extraction, and the line flushed after each draw. A 5 mL blood sample was collected in a vacutainer serum separator tube. It was not possible to perform blood collection exactly at the same time of day (e.g., 8 AM) for all participants; however, for each participant, samples were collected consistently at the same time of day across timepoints. Participants were asked to refrain from eating, smoking or drinking caffeinated drinks three hours before the blood extraction procedure. Following collection, blood samples were clotted for one hour, resting at room temperature, followed by 30 minutes at ∼4⁰C, and then centrifuged at 2200g for 15 minutes. The resulting serum was then aliquoted into 250μL cryovials and stored in a −80⁰C freezer. Serum samples were quantified in duplicate using Luminex assays for IL-6, IL-8, TNF-α, and IL-1β (Milliplex HSTCMAG-28SK-04, Millipore Sigma) as well as CRP (Milliplex HCVD3-MAG, Millipore Sigma) according to the manufacturer’s instructions. When concentration values were below the threshold of detection, the lowest detectable value was imputed.

### 2.5 Statistical analyses

Participant demographic statistics are summarized in frequencies (n, %) and means (standard deviation -SD-) or medians (interquartile range -IQR-) for normally and non-normally distributed data, respectively. Changes in the concentration of CRP, IL-6, IL-8, TNF-α, IL-1β, and IL-10 were analyzed using an intention-to-treat approach via repeated linear mixed models (LMMs). A per-protocol analysis including only participants with complete data at all time points was also performed. Outcome data were log-transformed to address non-normality of residuals. Furthermore, sensitivity analyses on non-log-transformed data were performed to assess the effect of normalization. Fixed effects in the LMMs included Group (CE, SC), Time (T0, T1), and their interaction (Group x Time). The covariates biological sex and age were entered as fixed factors given their established effects on inflammatory marker concentrations (Martinez de Toda et al., 2023). We also included participant-specific random intercepts and random slopes for time with an autoregressive correlation structure (AR1) to accommodate serial dependence. Missing outcome data were considered at random and handled via restricted maximum likelihood (REML). Beta (β) coefficients with their respective 95% confidence intervals (CIs) are reported. Pairwise comparisons between groups and time points were explored with uncorrected pairwise *t*-tests and least squares mean estimates and mean differences (MDs) with 95% CIs are provided. Given the exploratory nature of the analyses and the number of biomarkers, multiple comparison correction procedures were not applied. Exploratory *Spearman’s* correlations (*r^2^*) were used to explore associations among inflammatory mediators both at baseline and pre-post intervention (Δ=concentration T1-concentration T0). The same approach was used to explore associations between T0 to T1 changes in the concentration of inflammatory mediators and changes in cardio-respiratory fitness (VO_2_ peak), cognition (MoCA), and the degree of neurological (NIHSS) and motor (Fugl-Meyer) deficit. All analyses were conducted using JMP 19 pro with a significance level set at <0.05.

## 3. Results

Seventy-six participants were recruited in the original RCT (De Las Heras et al., 2025), but only 71 patients with blood samples available for the intention to treat analysis of the inflammatory mediators were included in this study (**Figure 1**). All participants assigned to the CE group who completed the study attended 24 training sessions and, despite some dropouts unrelated to participation (**Figure 1**), no serious adverse events were registered during training. **Table 1** presents the characteristics and other relevant clinical information of these 71 patients at baseline. During the duration of the study patients in the CE group received, on average, 8.3(8.06), 11.68(8.32), and 5.10(8.71) sessions of physio, occupational and speech therapy, respectively. Similarly, patients in the SC group received, on average, 6(5.23), 7.23(6.03), and 2.38(5.44) sessions of physio, occupational and speech therapy, respectively. The levels of physical activity outside the study assessed with the PASIPD were 8.18(5.41) and 9.89(7.19) METs hour/day for the CE and SC group, respectively. The SC group improved VO_2_peak by 0.29(2.52) mL.Kg^-1^.min^-1^ and the SC group by 4.49(3.24) mL.Kg^-1^.min^-1^, which is clinically relevant (Pandey et al., 2016).

**Figure 1.**
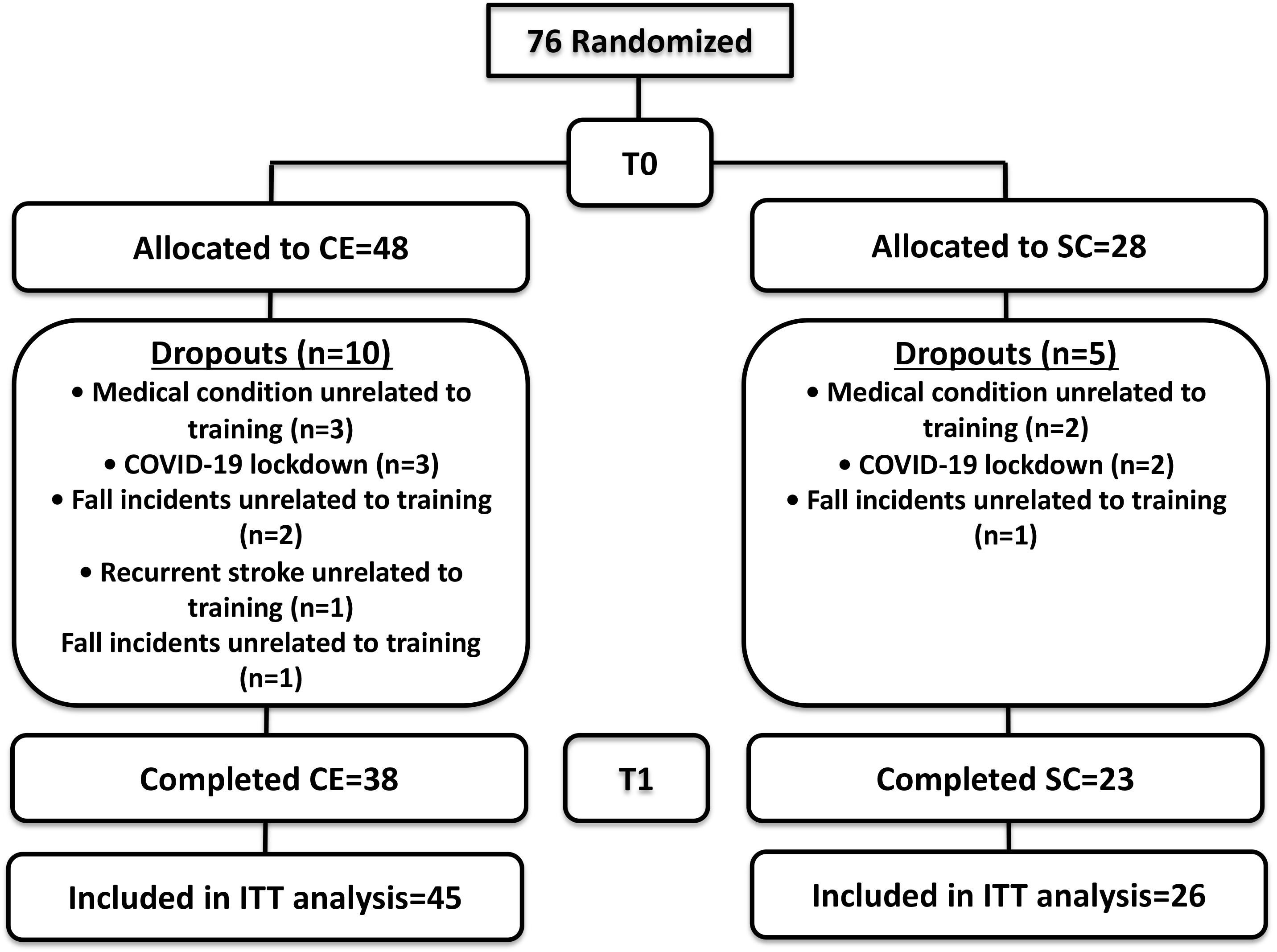
Participant flow diagram. Seventy-six participants were allocated into the CE (n=48) and SC (n=28) groups, respectively. Data from 71 participants were included in the intention-to-treat analysis.

**Table 1.** Participants baseline demographics.

|  | **CE (n=45)** | **SC (n=26)** |
| --- | --- | --- |
| Age, years | 61.77 (10.65) | 65.70 (8.89) |
| Females/Males | 11/34 | 10/16 |
| BMI, $\text{kg/m}^2$ | 27.62 (4.32) | 26.62 (3.97) |
| NIHSS (median, [IQR]) | 1 [4] | 2 [2] |
| MoCA | 24.31 (4.88) | 23.35 (8.47) |
| Fugl Meyer Total Score | 55.87 (10.48) | 58.88 (8.48) |
| Time post-stroke, days | 67.91 (20.10) | 59.96 (24.47) |
| Medications (n) | 5.13 (2.52) | 5.4 (2.20) |
| Classification (%) |  |  |
| AC | 58 | 53 |
| ACE | 33 | 44 |
| AP | 49 | 61 |
| BB | 38 | 27 |
| PSY | 34 | 26 |
| STA | 78 | 100 |
| Age-adjusted Charlson Index (median, [IQR]) | 4 [1] | 5 [3] |
| Resting SBP, mmHg | 128 (15) | 127 (17) |
| Resting DBP, mmHg | 76 (12) | 76 (10) |
| $\dot{V}O_{2peak}$, mL/kg/min | 17.97 (5.63) | 17.87 (5.09) |
| PASIPD, METs hour/day | 7.45 (5.08) | 9.51 (6.59) |

Values are means (SD) unless otherwise noted. AC: anticoagulant; ACE: angiotensin-converting Enzyme; AP: antiplatelet; BB: beta-blocker; BMI: body mass index; CCI: Charlson comorbidity index; DBP: diastolic blood pressure; MET: Metabolic equivalent; MoCA: Montreal Cognitive Assessment; NIHSS: National Institute of Health Stroke Scale; PASIPD: Physical Activity Scale for People with Disabilities; PSY: psychoactive; SBP: systolic blood pressure; STA: statin; $\dot{V}O_{2peak}$ : peak oxygen uptake.

### 3.1 Inflammatory mediators

#### 3.1.1 Baseline concentrations and associations between inflammatory mediators

Raw baseline concentration values of all inflammatory mediators are reported in supplementary tables (**Table S1**). Most baseline concentration values, including IL-10 were above commonly reported ranges for non-disabled individuals (**Figure S1 a-f**). Concentration values were above the threshold of detection in all mediators except for IL-1β, which had 28 participants with lowest detectable values imputed. This was not unexpected as IL-1β concentrations can usually be under the lower limit of detection (Kleiner et al., 2013) and IL-1β peaks very early after stroke and then it rapidly declines to baseline levels (Iordache et al., 2025). The results of the exploratory analyses investigating associations between the concentration of inflammatory mediators at baseline and pre-post intervention changes are provided in supplementary tables (**Table S4** and **S5**, respectively). In short, in the CE group, increases in IL-6 were associated with increases in IL-8, IL-10 and TNF-α. In contrast, in the SC group increases in IL-10 were associated with increases in IL-6, IL-8, TNF-α and IL-1β, and increases in IL-8 with increases in TNF-α. Hence, despite being non-inflammatory, IL-10 appeared to increase in concert with pro-inflammatory precursors.

#### 3.1.2 Differences between CE and SC in inflammatory mediators after the intervention

The fixed effects from the adjusted LMMs for the intention to treat analysis are shown in **Table 3**. Group × Time interactions were statistically significant for IL-6, IL-8, and TNF-α. Interactions, in contrast, were not significant for CRP, IL-10, or IL-1β. The results of the sensitivity analyses using non-log-transformed data aligned with the results reported here with one exception; differences between groups for the CRP Group × Time interaction became statistically significant, with the SC group showing increased concentrations at T1 in comparison to the CE group (β=13.896, 95% CI=62.260, 27.729 p=0.049) (**Figure S1 a**). The discrepancy between the results of non-log-transformed and log-transformed analyses in CRP concentration is explained by the influential effect of two observations in the SC that were 10 and 15-fold above the mean group at T1. The influential effect of these two observations on the LMM decreased when data were log-transformed. More details about each LMM and the effects of each variable on the models of the intention to treat analysis are provided in supplementary tables (**Table S2**).

Adjusted least square mean estimate plots showing the trajectory of each inflammatory mediator are provided in **Figure 2a-f**. Pairwise *t*-tests showed no significant between-group differences at T0. The CE group only showed a significant within-group change -reduction-from T0 to T1 in TNF-α (MD=0.06, 95% CI=0.02,0.11; p=0.003). In contrast, the SC group demonstrated significant increases in IL-6 (MD=−0.40, 95% CI=−0.74,-0.07, p=0.019), IL-8 (MD=−0.14, 95% CI=−0.27,-0.02, p=0.024), and IL-10 (MD=−0.24, 95% CI=−0.46, −0.02, p=0.031). Between-group pairwise comparisons at T1 indicated that the values for CRP (MD=−0.74, 95% CI=−1.47, −0.01, p=0.045), IL-8 (MD=−0.45, 95% CI=−0.84,-0.07, p=0.022), and TNF-α (MD=−0.33., 95% CI −0.61,-0.05, p=0.021) were significantly higher in the SC group than in the CE group. Regardless of the intervention, IL-1β remained relatively stable and showed no significant group differences at either timepoint. Taken together, pairwise *t*-test results aligned with the LMMs findings (**Table 2**), indicating that the concentration of inflammatory mediators tended to rise in the SC group but remained stable or declined in the CE group, thus producing differences between groups at T1. To allow comparison with other studies, adjusted least square non-log-transformed mean estimates with normative data reference ranges published previously are provided in supplementary files (**Figure S1a-f**).

**Figure 2.**
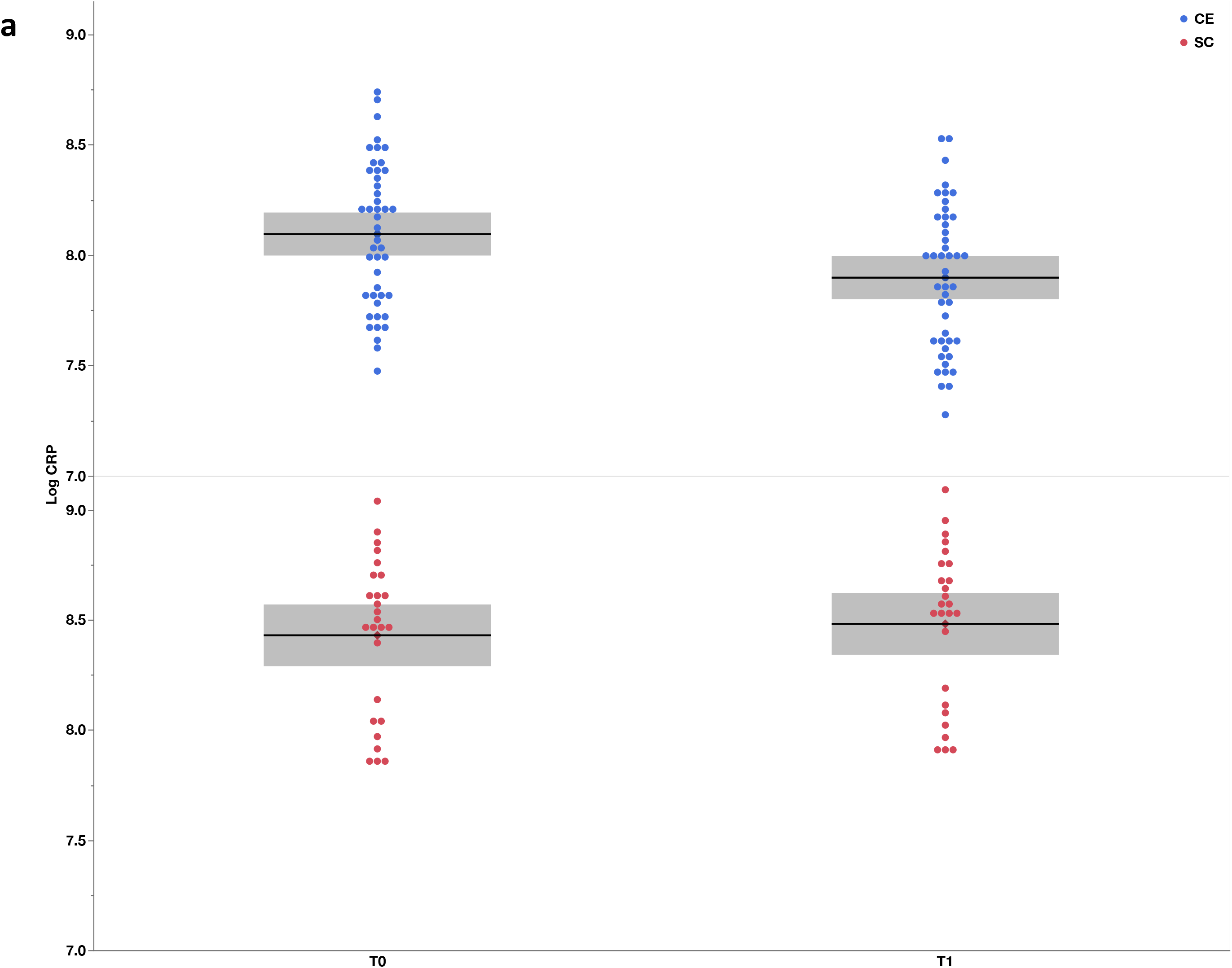
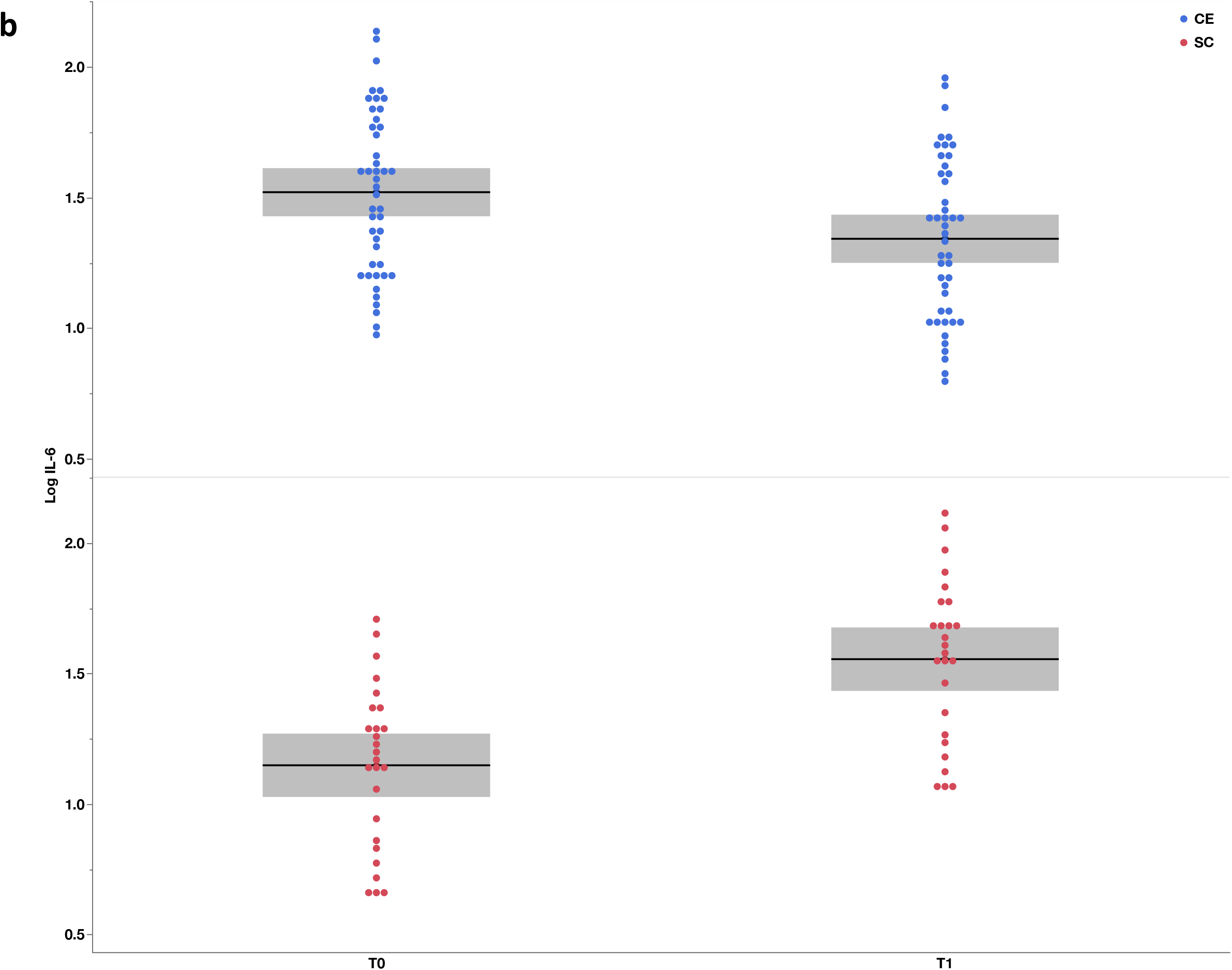
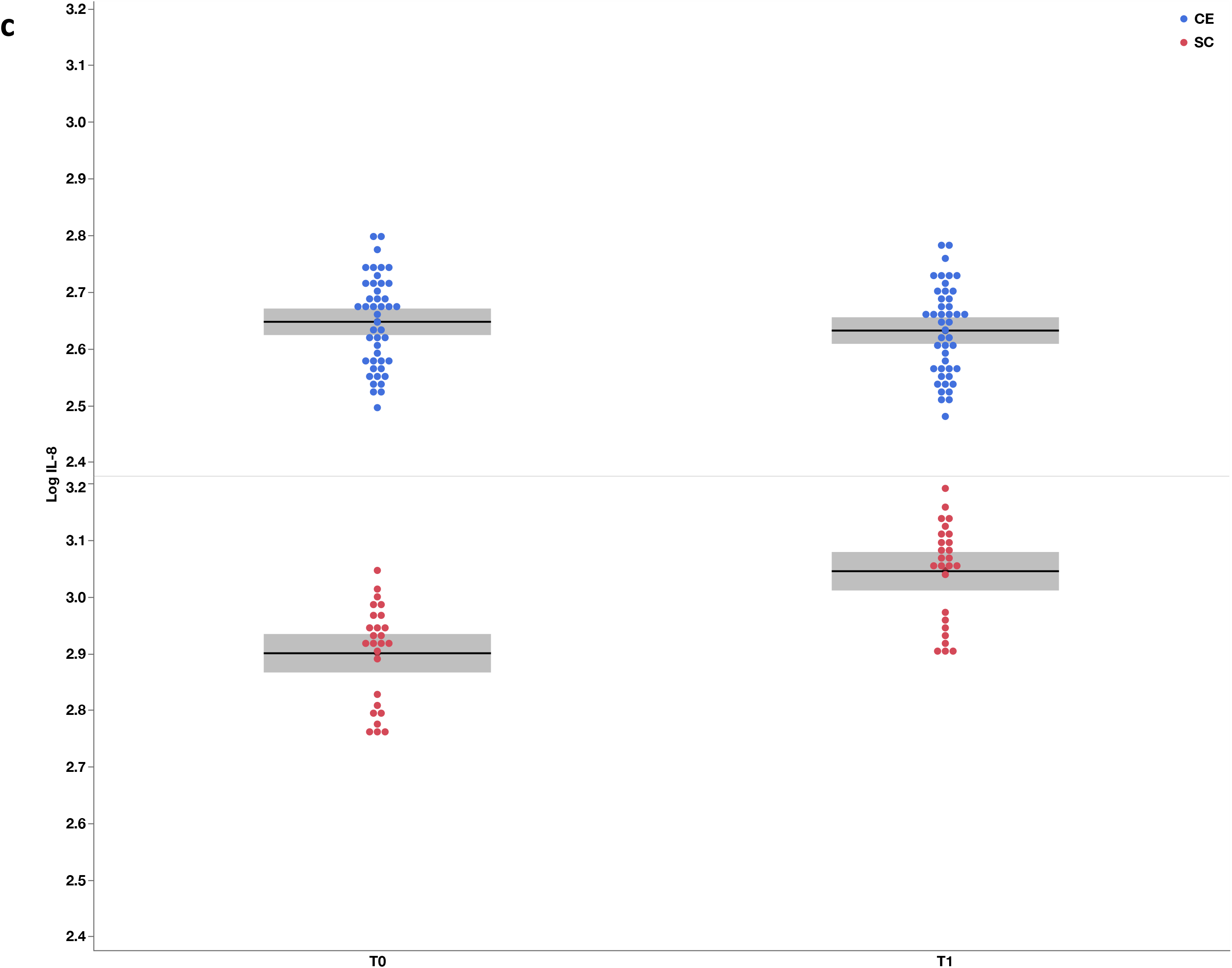
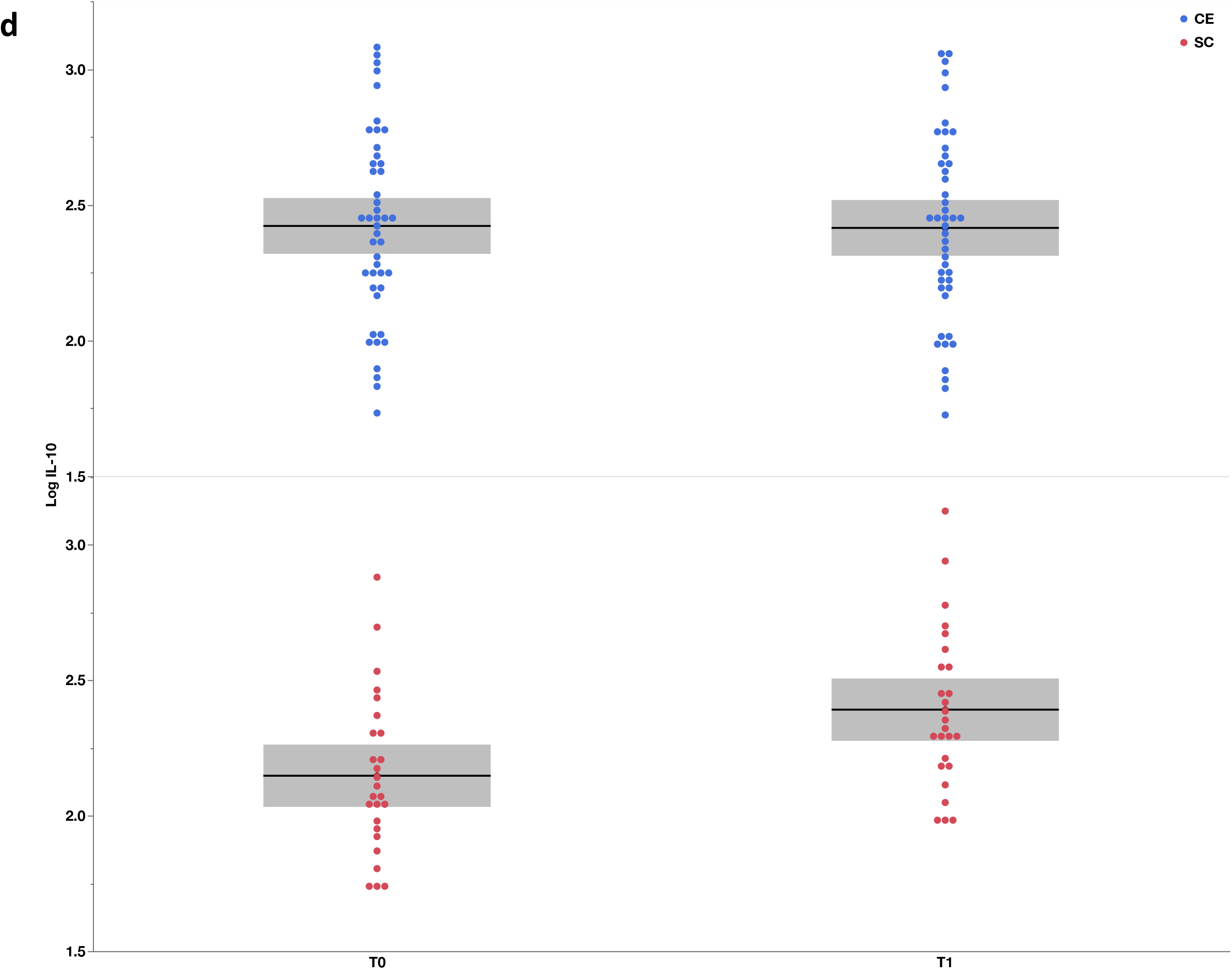
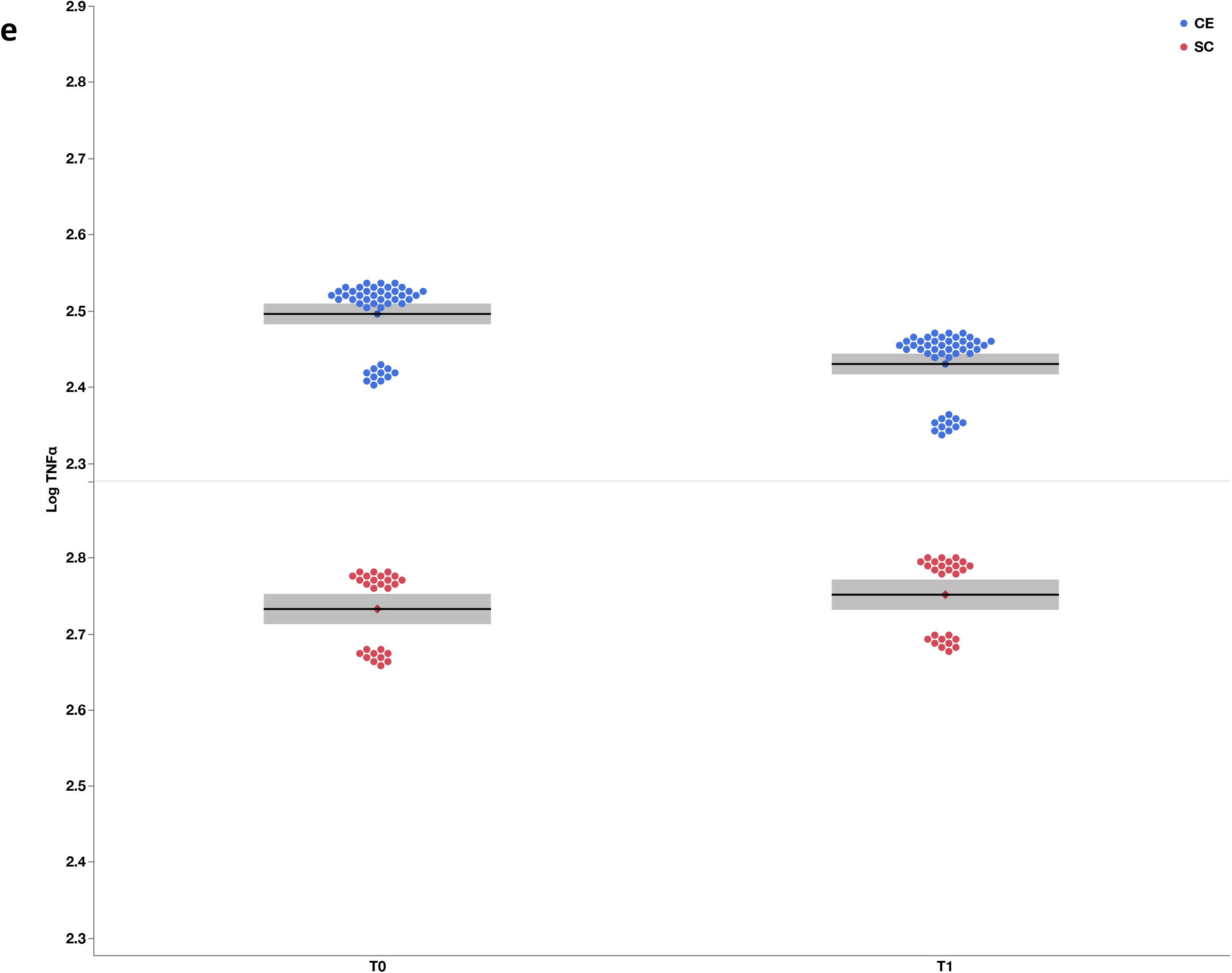
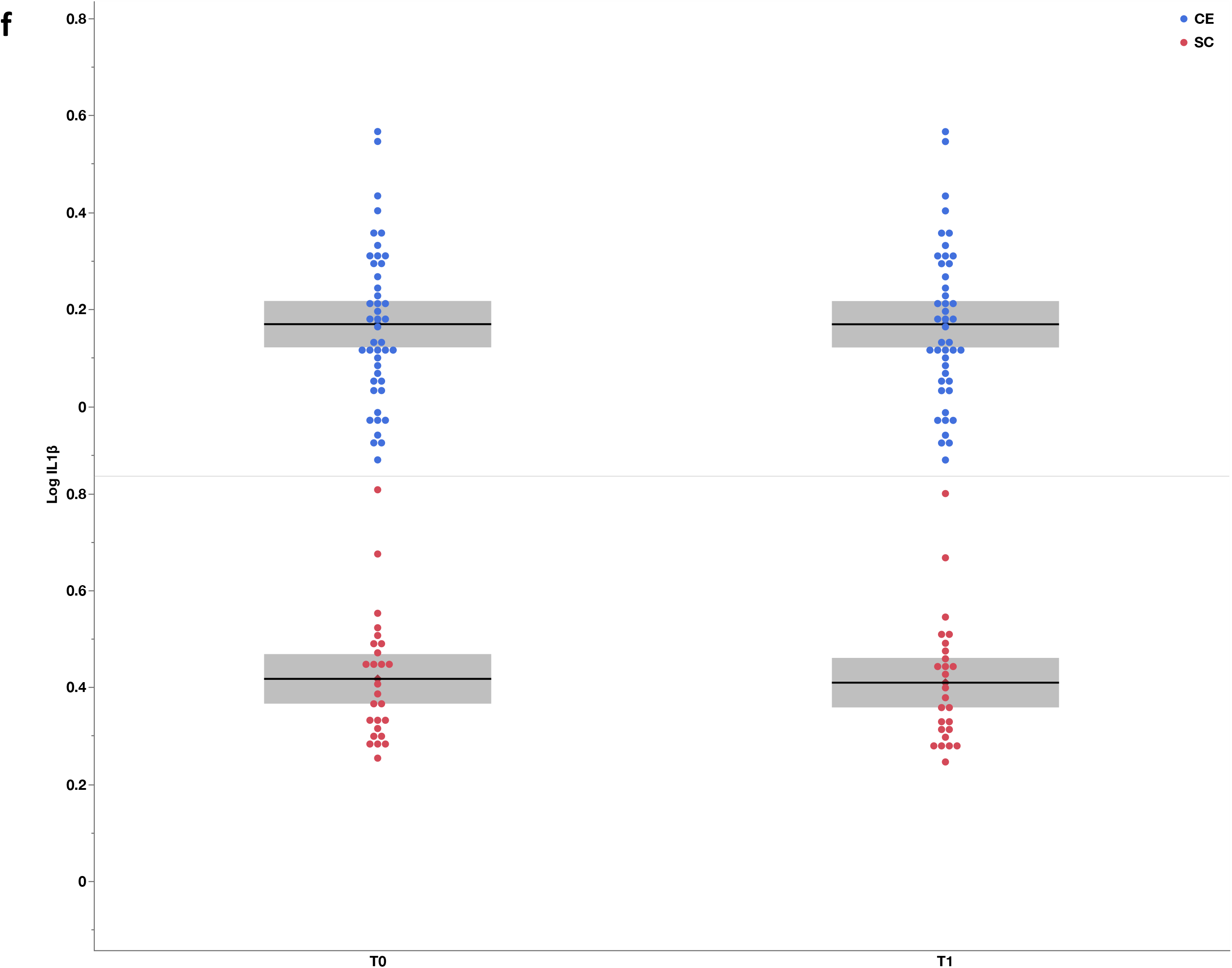
**a-f.** Least square mean estimates plots showing changes in the concentration of the inflammatory mediators CRP, IL-6, IL-8, IL-10, TNF-α and IL-1β for CE and SC from T0 to T1. Values are log-transformed predicted mean estimates with 95% CIs.

**Table 2.**
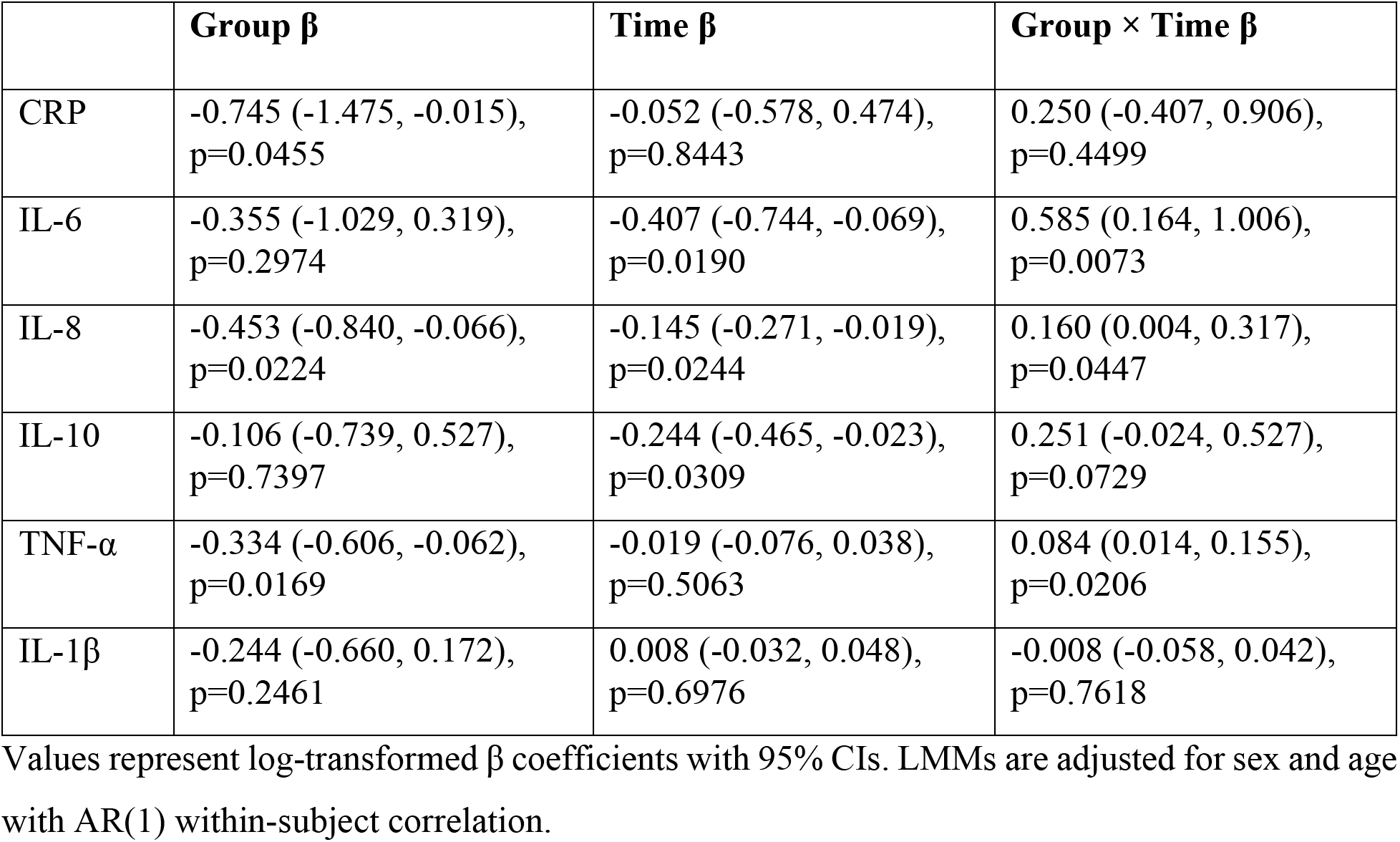
Fixed effects parameter estimates (β) from the LMMs adjusted for age and sex.

| | Group $\beta$ | Time $\beta$ | Group × Time $\beta$ |
| --- | --- | --- | --- |
| CRP | -0.745 (-1.475, -0.015),<br>$p=0.0455$ | -0.052 (-0.578, 0.474),<br>$p=0.8443$ | 0.250 (-0.407, 0.906),<br>$p=0.4499$ |
| IL-6 | -0.355 (-1.029, 0.319),<br>$p=0.2974$ | -0.407 (-0.744, -0.069),<br>$p=0.0190$ | 0.585 (0.164, 1.006),<br>$p=0.0073$ |
| IL-8 | -0.453 (-0.840, -0.066),<br>$p=0.0224$ | -0.145 (-0.271, -0.019),<br>$p=0.0244$ | 0.160 (0.004, 0.317),<br>$p=0.0447$ |
| IL-10 | -0.106 (-0.739, 0.527),<br>$p=0.7397$ | -0.244 (-0.465, -0.023),<br>$p=0.0309$ | 0.251 (-0.024, 0.527),<br>$p=0.0729$ |
| TNF-α | -0.334 (-0.606, -0.062),<br>$p=0.0169$ | -0.019 (-0.076, 0.038),<br>$p=0.5063$ | 0.084 (0.014, 0.155),<br>$p=0.0206$ |
| IL-1β | -0.244 (-0.660, 0.172),<br>$p=0.2461$ | 0.008 (-0.032, 0.048),<br>$p=0.6976$ | -0.008 (-0.058, 0.042),<br>$p=0.7618$ |

Values represent log-transformed $\beta$ coefficients with 95% CIs. LMMs are adjusted for sex and age with AR(1) within-subject correlation.

The results of the per-protocol analysis, which are shown in supplementary **Table S3**, coincided with the results of the intention to treat analysis, with Group × Time interactions for IL-6 (β=0.590, 95% CI=0.167, 1.014, p=0.0071), IL-8 (β=0.165, 95% CI=−0.005, 0.325, p=0.043), and TNF-α (β=0.083, 95% CI=0.014, 0.152, p=0.020). The rest of inflammatory mediators showed the same trajectories of the intention to treat analysis and differences between groups did not reach statistical significance.

#### 3.1.3 Associations between changes in inflammatory mediators and clinical measures

Detailed results of the analyses exploring associations between pre-post intervention changes in the concentration of inflammatory mediators and changes in clinical measures are shown in supplementary **Table S6**. The CE group showed negative associations between motor deficit (Fugl-Meyer) and the concentration of IL-6 (*r^2^*=−0.52, p=0.039) and IL-10 (*r^2^*=−0.53, p=0.018). In other words, participants with larger reductions in these two inflammatory mediators after training tended to show greater reductions in motor deficits. In the CE group we also found positive association between IL-1β and cardio-respiratory fitness (VO_2peak_) but, due to the large number of imputations for this biomarker, this result should be interpreted very cautiously. The analyses of the SC group did not reveal any significant association between any inflammatory mediator and clinical measure.

## 4. Discussion

Ischemic stroke triggers a cascade characterized by acute elevations in pro-inflammatory mediators (CRP, IL-6, IL-8, TNF-α, IL-1β) and a delayed counter-regulatory rise in anti-inflammatory mediators (IL-10) (Iordache et al., 2025). In short, after vessel occlusion, microglia and infiltrating immune cells release IL-1β and TNF-α, promoting blood–brain barrier disruption, leukocyte adhesion, and neuronal apoptosis (Shi et al., 2019). Driven by IL-1β and TNF-α signaling, IL-6 further expands this initial cascade by increasing leukocyte recruitment and stimulating hepatic production of CRP, a biomarker of systemic inflammation that peaks 48–72 hours post-stroke (Zhu et al., 2022). Concomitantly, IL-8 released by activated glial and endothelial cells accelerates neutrophil chemotaxis, exacerbating tissue damage (Zhu et al., 2022). IL-10 emerges during the late hyperacute phase (24 hours post-stroke), attenuating the production of pro-inflammatory mediators (IL-6, TNF-α, IL-1β) (Zhu et al., 2022), oxidative stress, and contributing to neural recovery (Schmidt-Pogoda et al., 2025).

Inflammation is a promising therapeutic target for stroke rehabilitation (Couch et al., 2022) and secondary prevention (Zietz et al., 2024). Recent pre-clinical and early clinical biomarker-guided immunomodulation trials have explored the possibility of inhibiting the action of these mediators (e.g., IL-6, IL-1β) to suppress inflammation and reduce secondary neural damage post-stroke and disability (Brough et al., 2015; Iordache et al., 2025). Here, we investigated the capacity of CE, a widely accessible behavioral intervention with ample effects on multiple inflammatory pathways (Muller et al., 2021). Specifically, we hypothesized that CE would reduce and increase the concentration of pro-inflammatory and anti-inflammatory mediators, respectively (Poorhabibi et al., 2025; Tayebi et al., 2025). While our results partially support our initial hypothesis, the study provided three novel findings. First, rather than lowering the inflammatory response, our data suggest that CE maintains inflammation levels stable. When CE is not implemented, like in the SC group, inflammation can be exacerbated (**Figure 1**). Second, intersubject variability in the response to CE is substantial and the anti-inflammatory effect differs substantially across mediators. Third, in this early subacute phase of recovery, CE does not seem to increase IL-10, an important anti-inflammatory mediator, whose reduced concentration has been associated with negative stroke outcome (Sun et al., 2021).

None of our participants reported infections, were diagnosed with new comorbidities or had changes in medications during the study that could have affected inflammation. Since the concentration of inflammatory mediators (CRP, IL-6 and TNF-α) tends to follow a natural declining trajectory after stroke, the increase in inflammation during the intervention period in the SC group was rather unexpected. It should be noted, however, that the typical declining inflammatory trajectory has been established through studies using blood samples at admission or some hours after the stroke event as baseline. As it is precisely during these first hour post-stroke where the acute inflammatory response tends to reach its highest levels (Carmichael et al., 2026; Sandvig et al., 2023), a declining trajectory is therefore expected. However, there is much less information regarding potential modulations in the trajectory of these inflammatory mediators during the transition from early to late sub-acute stages of stroke recovery like the ones covered in our study. In fact, recent studies have shown that transient increases in inflammation during subacute periods of recovery can co-exist within the characteristic long-term declining trend (Kirzinger et al., 2021). More studies are needed to characterize inflammation in these subacute stages and determine how reducing it could impact different aspects of long-term stroke recovery (Carmichael et al., 2026; Sandvig et al., 2023) and recurrence (McCabe et al., 2024).

Few studies have investigated the impact of exercise-based interventions on inflammatory mediators post-stroke and results have been inconsistent (Bitencourt et al., 2025; Couch et al., 2022). Furthermore, most studies had relatively small sample sizes, included chronic patients, used mild to moderate exercise intensities and focused exclusively on CRP, IL-6 and TNF-α (Bitencourt et al., 2025). One study (n=63) found that 12 weeks of home-based high intensity interval CE performed five times per week downregulated IL-6 levels among patients with recent (21 days post-stroke) lacunar stroke. However, it is important to note that differences with the control group were not statistically significant (Krawcyk et al., 2019). Another study (n=30) found reductions in IL-6 after four weeks of CE performed five days per week at moderate intensity in patients with chronic stroke (Hsu et al., 2019). In contrast, one large study (n=200) failed to show changes in CRP, IL-6 or TNF-α following four weeks of very light-intensity CE performed five days per week in very early subacute patients (17-40 days post-stroke) (Kirzinger et al., 2021). Similarly, a study in which chronic stroke patients (n=39) performed six months of CE three times per week at a moderate-to-vigorous intensity also failed to demonstrate significant changes in CRP and IL-6 (Serra et al., 2022).

Our study provides new evidence demonstrating that, when introduced in the early subacute phase of stroke recovery, moderate-to-vigorous CE mitigates increases in the concentration of pro-inflammatory mediators IL-6, IL-8 and TNF-α. Compared to the SC group, the CE group also showed declines in CRP, but differences became significant only in analyses using non-log-transformed data. CE either maintained within or reduced the concentration of inflammatory mediators in relation to normative ranges associated with no inflammation (**Figure S1a-f**). However, since associations between pre-post intervention changes in the concentration of inflammatory mediators and changes in clinical measures were negligible (**Table S6)**, to what extent the mediator selective anti-inflammatory effect of CE shown in this study is enough to bring about clinical benefit for these patients is unclear.

Given the heterogeneity among studies in terms of patients’ characteristics, exercise and control interventions as well as blood analysis methodology, the exact reasons for the discrepancies between the results of our study and previous studies are difficult to ascertain. The study that perhaps offers the best direct comparison is the study by Krawcyk et al., who found decreases in IL-6, only in the CE group - no differences with the control group - after 12 weeks of home-based high intensity interval CE (Krawcyk et al., 2019). Our cohort was more responsive to the effects of CE because we did find significant between group differences in IL-6 and TNF-α change. Differences between studies could be potentially explained by the fact that the study by Krawcyk et al., only included patients with mild (lacunar) strokes who had, at baseline, 7-10-fold lower mean concentrations of IL-6 (1.10 pg/mL) and TNF-α (2.22 pg/mL) than the participants of our study (IL-6=10.26 pg/mL and TNF-α=15.22 pg/mL).

Another study that investigated the effects of CE on early subacute patients was the study by Kirzinger et al., who showed no effects of 25 mins of bodyweight supported treadmill CE performed five times weekly for four weeks on CRP, IL-6 or TNF-α concentration (Kirzinger et al., 2021). This study is relevant because, besides looking at the effects of CE, researchers also measured the concentration of inflammatory biomarkers four weeks, and three and six months after the stroke event, thus providing trajectories in sub-acute stages of recovery. While in this case the mean baseline concentration levels of CRP (12.04 mg/mL), IL-6 (6.34 pg/mL) and TNF-α (9.34 pg/mL) of this study were more aligned with our mean values (CRP=8.14 mg/mL; IL-6=10.26 pg/mL; TNF-α=15.22 pg/mL), the study cannot be used for direct comparison because their patients were substantially more impaired (Median [IQR] NIHSS score= 8[7]). More importantly, as acknowledged by the authors of the study, the duration (four weeks) and intensity (very mild to mild) of the CE intervention was probably insufficient to reduce inflammation.

Based on all these studies one could speculate that early subacute patients with higher levels of inflammation, exposed to vigorous long-term CE programs will show the largest improvements in inflammation. However, we do not have enough evidence to date to identify a single driver of the anti-inflammatory effect of CE post-stroke and the reason why studies show divergent results. Larger studies including patients with broader levels of disability post-stroke and with better stratification are needed to determine which patients can benefit the most from CE, which phases of stroke recovery are more sensitive to its effects and, importantly, which exercise parameters (e.g., frequency, intensity, duration and type) drive the reduction in inflammation.

IL-10 is a key anti-inflammatory cytokine that plays a protective role in stroke recovery by regulating the immune response following ischemic brain injury (Xiao et al., 2025). After a stroke, excessive inflammation can contribute to secondary neuronal damage and infarct expansion and IL-10 helps limit this process by suppressing pro-inflammatory cytokines and reducing harmful immune cell activity (Piepke et al., 2021). The lack of increases in the concentration of IL-10 in response to CE was unexpected. CE can increase IL-10 in older individuals (Tayebi et al., 2025) and recent animal studies provide evidence that this anti-inflammatory cytokine is upregulated with CE, contributing to stroke recovery by reducing neuronal hyperexcitability (Schmidt-Pogoda et al., 2025). However, while some post-stroke human trials investigating the effects of CE on IL-10 are underway (Oliveira et al., 2019), for now there is not robust evidence in support of using this intervention to upregulate this cytokine in patients with stroke (da Cunha et al., 2024). The reason for the lack of IL-10 increase after CE is unclear but one possible explanation could be that our cohort (Median[IQR]=8.83 [13.46]), and especially the CE group (Median[IQR]=10.46 [16.13]) already had high concentration levels (Kleiner et al., 2013) that were well above the levels previously reported post-stroke (Sun et al., 2021).

Importantly, exploratory correlational analyses revealed positive associations between changes in IL-10 and IL-6 in the CE group (*r^2^*=0.59) (**Table S4**). Since one of the mechanisms through which IL-10 reduces inflammation is through the inhibition of IL-6 (Zhu et al., 2022), we were expecting this correlation to have a negative sign (e.g., higher IL-10 concentration associated with lower IL-6 concentration) (Sun et al., 2021). However, due to their different time-courses, it is possible for pro- and anti-inflammatory mediators like IL-10 and IL-6 to rise simultaneously and overlap in some periods of stroke (Iordache et al., 2025). The fact that reductions in both IL-10 and IL-6 concentration were associated with improvements in motor function in the CE group reinforce this view. Given the relevance of IL-10 in mitigating inflammation and improving neural repair post-stroke (Zhu et al., 2022), more studies are needed to confirm these results and to determine the capacity of CE to upregulate this anti-inflammatory cytokine in response to brain ischemia (Oliveira et al., 2019).

## 5. Limitations

This secondary analysis of an RCT presents several limitations that should be considered when interpreting its findings. First, participants in both groups continued to receive usual rehabilitation and, although differences were not statistically significant, the CE group attended numerically more physiotherapy and occupational therapy sessions than the SC group. Given their low intensity, it is unlikely that these therapies had any effect on inflammation, but we cannot fully exclude residual confounding by adjunctive rehabilitation. Second, six biomarkers were analyzed without correction for multiple comparisons. The findings for IL-6, IL-8 and TNF-α are therefore exploratory and hypothesis-generating, carry an increased risk of type I error, and should be confirmed in adequately powered studies. Third although multiple inflammatory mediators were examined, we did not assess composite inflammatory indices or additional biomarkers (e.g., IL-4, IFN-γ, adhesion molecules) that could provide a broader characterization of the systemic inflammatory response post-stroke (Lai et al., 2019). Fourth, while blood samples were collected at consistent times within each participant, diurnal variability could not be fully controlled across participants (Zhou et al., 2010). Fifth, the intervention lasted eight weeks, but we did not include follow-up assessments; therefore, it remains unclear whether the anti-inflammatory effects of CE persisted beyond the intervention period or translate into long-term clinical or functional benefits. Our findings therefore cannot support conclusions about sustained effects on stroke recovery or recurrence. Sixth, the associations between changes in biomarkers and clinical measures were exploratory and do not establish a mechanism. Seventh, although the sample size was larger than that of most previous inflammatory biomarker-exercise studies in stroke (Bitencourt et al., 2025), sample size was estimated to detect differences in the main outcomes of the RCT only (NCT05076747). In addition, despite the use of LMMs, which are especially well suited for handling imbalanced between-group data while preserving power, the use of a 2:1 randomization ratio could have affected power. Eight, intersubject variability remained high, limiting our ability to explore dose–response relationships or stratify effects based on stroke severity, lesion characteristics, or baseline inflammation. Finally, given the elevated number of IL-1β samples with levels below the threshold of detection (Kleiner et al., 2013), the results pertaining to this cytokine should be interpreted very cautiously. Blood biomarkers provide only an indirect estimate of neuroinflammation, and we did not include neuroimaging or cerebrospinal fluid measures that could strengthen mechanistic inferences regarding brain-specific inflammatory processes.

## Conclusions

Eight weeks of moderate-to-vigorous CE introduced in the early subacute phase of stroke recovery mitigated increases in some key pro-inflammatory mediators, including IL-6, IL-8, and TNF-α. These findings suggest that CE may help maintain, rather than merely reduce, the exaggerated inflammatory response characteristic of the early post-stroke period. However, the anti-inflammatory effect was not uniform across mediators, and no decreases in CRP or increases in IL-10 concentrations were observed. This highlights the complexity of the inflammatory cascade and the need to understand differential biomarker responsiveness.

Given the established links between post-stroke inflammation, neural repair, and secondary stroke risk, CE represents a promising, accessible, and low-cost intervention to improve biological conditions for recovery. Future studies with larger cohorts, biomarker stratification, and long-term follow-up are needed to confirm these findings, identify responders, determine optimal exercise parameters, and examine whether early modulation of inflammation through exercise, especially in early subacute stages of recovery, translates into improved long-term functional outcomes and reduced recurrence risk.

## Data Availability

All data produced in the present study are available upon reasonable request to the authors

## Acknowledgements

The authors thank Sandra Da Cal and Gloria Gabrielle Delgado for their excellent work in performing the Luminex assays and Olfa Debbeche from the CRCHUM biosafety platform for access to the Luminex reader.

## Supplementary Tables

**Table S1.**
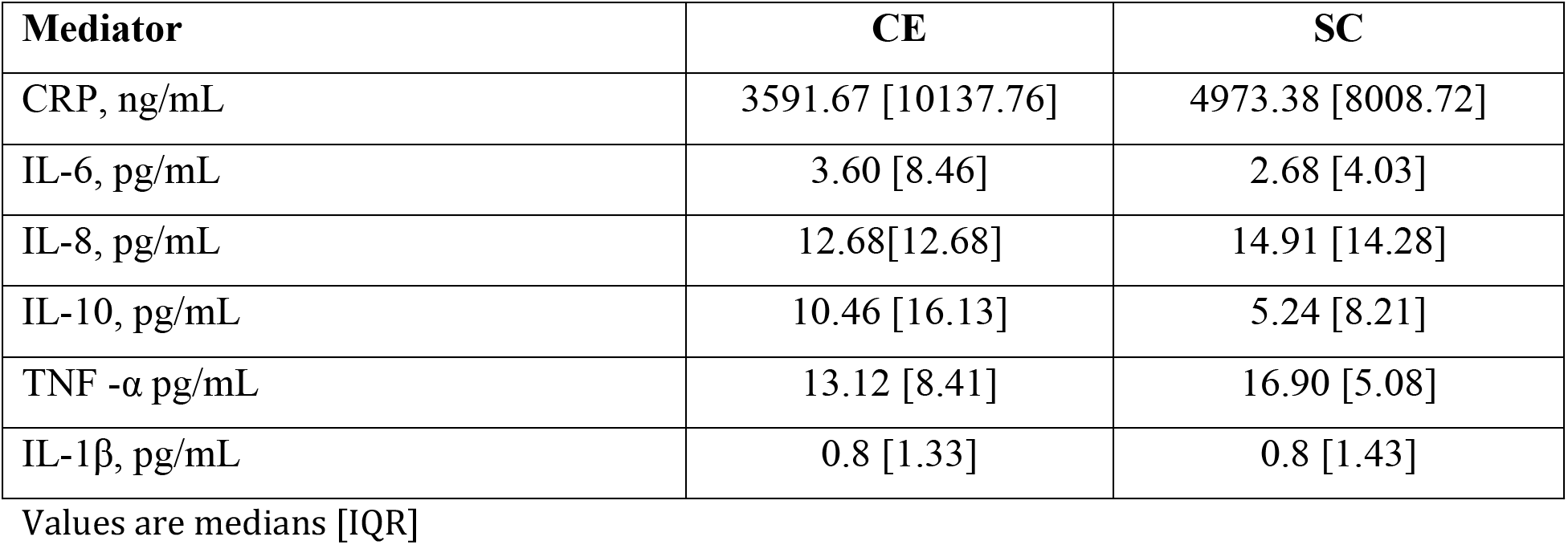
Baseline values of inflammatory mediators for cardiovascular exercise (CE) and standard care (SC) groups.

| Mediator | CE | SC |
| --- | --- | --- |
| CRP, ng/mL | 3591.67 [10137.76] | 4973.38 [8008.72] |
| IL-6, pg/mL | 3.60 [8.46] | 2.68 [4.03] |
| IL-8, pg/mL | 12.68[12.68] | 14.91 [14.28] |
| IL-10, pg/mL | 10.46 [16.13] | 5.24 [8.21] |
| TNF -α pg/mL | 13.12 [8.41] | 16.90 [5.08] |
| IL-1β, pg/mL | 0.8 [1.33] | 0.8 [1.43] |

Values are medians [IQR]

**Table S2.** Fixed effects parameters (intention to treat analysis)

| Parameter | $\beta$ | Std Error | DF Den | 95% CI | p value |
| --- | --- | --- | --- | --- | --- |
| **CRP** |  |  |  |  |  |
| Intercept | 10.0928 | 1.041 | 67.4 | 8.0151, 12.1704 | <0.0001 |
| Group | -0.7454 | 0.3681 | 104.4 | -1.4753, -0.0154 | 0.0455 |
| Time | -0.0519 | 0.263 | 60.2 | -0.5779, 0.4742 | 0.8443 |
| Group×Time | 0.2496 | 0.3282 | 60.0 | -0.4069, 0.9061 | 0.4499 |
| Age | -0.028 | 0.0148 | 65.7 | -0.0575, 0.0015 | 0.0627 |
| Sex | 0.3695 | 0.3293 | 69.1 | -0.2874, 1.0263 | 0.2657 |
| **IL-6** |  |  |  |  |  |
| Intercept | 3.2746 | 1.046 | 65.9 | 1.1863, 5.363 | 0.0026 |
| Group | -0.3553 | 0.3389 | 85.3 | -1.0291, 0.3185 | 0.2974 |
| Time | -0.4067 | 0.1683 | 55.5 | -0.7439, -0.0695 | 0.0190 |
| Group×Time | 0.5848 | 0.2099 | 55.4 | 0.1642, 1.0055 | 0.0073 |
| Age | -0.0283 | 0.0149 | 65.2 | -0.0581, 0.0015 | 0.0620 |
| Sex | 0.2268 | 0.3293 | 66.8 | -0.4305, 0.8841 | 0.4934 |

| **IL-8** |  |  |  |  |  |
| --- | --- | --- | --- | --- | --- |
| Intercept | 3.423 | 0.6259 | 67.1 | 2.1737, 4.6723 | <0.0001 |
| Group | -0.4528 | 0.1942 | 75.1 | -0.8395, -0.066 | 0.0224 |
| Time | -0.145 | 0.0626 | 55.1 | -0.2705, -0.0195 | 0.0244 |
| Group×Time | 0.1605 | 0.0781 | 55.1 | 0.0039, 0.317 | 0.0447 |
| Age | -0.0066 | 0.0089 | 66.8 | -0.0245, 0.0112 | 0.4613 |
| Sex | 0.0933 | 0.1966 | 67.5 | -0.2991, 0.4857 | 0.6365 |
| **IL-10** |  |  |  |  |  |
| Intercept | 4.527 | 1.0191 | 66.5 | 2.4926, 6.5614 | <0.0001 |
| Group | -0.1059 | 0.3177 | 75.8 | -0.7386, 0.5267 | 0.7397 |
| Time | -0.244 | 0.1102 | 54.6 | -0.4648, -0.0232 | 0.0309 |
| Group×Time | 0.2513 | 0.1374 | 54.6 | -0.0241, 0.5267 | 0.0729 |
| Age | -0.0326 | 0.0145 | 66.2 | -0.0617, -0.0036 | 0.0284 |
| Sex | 0.0112 | 0.3202 | 66.9 | -0.628, 0.6503 | 0.9723 |
| **TNF-α** |  |  |  |  |  |
| Intercept | 2.6581 | 0.5628 | 46.6 | 1.5257, 3.7905 | <0.0001 |
| Group | -0.3339 | 0.1398 | 49.4 | -0.6149, -0.0529 | 0.0208 |
| Time | -0.0188 | 0.0279 | 36.3 | -0.0753, 0.0377 | 0.5037 |
| Group×Time | 0.0845 | 0.0347 | 35.8 | 0.0141, 0.1549 | 0.0200 |
| Age | 0.0004 | 0.0082 | 46.5 | -0.016, 0.0169 | 0.9572 |
| Sex | 0.1042 | 0.1432 | 46.7 | -0.1839, 0.3924 | 0.4703 |
| **IL-1β** |  |  |  |  |  |
| Intercept | 1.2382 | 0.6908 | 66.9 | -0.1408, 2.6171 | 0.0776 |
| Group | -0.2439 | 0.2085 | 67.6 | -0.6599, 0.1721 | 0.2461 |
| Time | 0.0078 | 0.02 | 54.0 | -0.0323, 0.048 | 0.6976 |
| Group×Time | -0.0076 | 0.025 | 54.0 | -0.0577, 0.0425 | 0.7618 |
| Age | -0.0102 | 0.0099 | 66.9 | -0.0299, 0.0095 | 0.3053 |
| Sex | -0.2582 | 0.2167 | 66.9 | -0.6908, 0.1743 | 0.2376 |

**Table S3.** Fixed effects parameters (per-protocol analysis)

| Parameter | $\beta$ | Std Error | DF Den | 95% CI | p value |
| --- | --- | --- | --- | --- | --- |
| **CRP** |  |  |  |  |  |
| Intercept | 9.5535363 | 1.1266783 | 53.5 | 7.2942175, 11.812855 | <0.0001 |
| Group | -0.675682 | 0.3975614 | 73.9 | -1.46786, 0.1164966 | 0.0934 |
| Time | -0.034922 | 0.2703739 | 54.0 | -0.576989, 0.507145 | 0.8977 |
| Group×Time | 0.1960446 | 0.3372155 | 54.0 | -0.480032, 0.872121 | 0.5634 |
| Age | -0.018662 | 0.016083 | 52.0 | -0.050935, 0.0136109 | 0.2512 |
| Sex | 0.3080263 | 0.3880717 | 52.0 | -0.470697, 1.0867491 | 0.4310 |
| **IL-6** |  |  |  |  |  |
| Intercept | 3.8680388 | 1.0651664 | 52.7 | 1.7312637, 6.0048138 | 0.0006 |
| Group | -0.365178 | 0.3576919 | 61.8 | -1.08023, 0.3498736 | 0.3113 |
| Time | -0.410823 | 0.1692044 | 54.0 | -0.750058, -0.071589 | 0.0185 |
| Group×Time | 0.5905329 | 0.211035 | 54.0 | 0.1674332, 1.0136326 | 0.0071 |
| Age | -0.038604 | 0.0152672 | 52.0 | -0.06924, -0.007968 | 0.0145 |
| Sex | 0.3133106 | 0.368387 | 52.0 | -0.425912, 1.0525332 | 0.3990 |
| **IL-8** |  |  |  |  |  |
| Intercept | 3.6133338 | 0.7387364 | 47.1 | 2.1272609, 5.0994068 | 0.0001 |
| Group | -0.418761 | 0.2235643 | 51.1 | -0.867567, 0.0300443 | 0.0668 |
| Time | -0.148301 | 0.06368 | 42.0 | -0.276815, -0.019787 | 0.0248 |
| Group×Time | 0.165089 | 0.0795211 | 49.9 | 0.00536, 0.324818 | 0.0431 |
| Age | -0.010707 | 0.0108542 | 46.8 | -0.032545, 0.0111311 | 0.3290 |
| Sex | 0.1421389 | 0.2336274 | 46.8 | -0.327912, 0.6121895 | 0.5459 |
| **IL-10** |  |  |  |  |  |
| Intercept | 5.2359721 | 1.0323072 | 52.3 | 3.1647775, 7.3071666 | <0.0001 |
| Group | -0.064249 | 0.3388757 | 56.5 | -0.742972, 0.6144749 | 0.8503 |
| Time | -0.249605 | 0.1104286 | 54.0 | -0.471001, -0.028209 | 0.0279 |
| Group×Time | 0.2631158 | 0.1377287 | 54.0 | -0.013014, 0.5392452 | 0.0614 |
| Age | -0.045759 | 0.0148219 | 52.0 | -0.075501, -0.016017 | 0.0032 |
| Sex | 0.1393133 | 0.3576415 | 52.0 | -0.578347, 0.8569734 | 0.6985 |
| **TNF-α** |  |  |  |  |  |
| Intercept | 2.4402889 | 0.4971418 | 52.1 | 1.4427381, 3.4378398 | <0.0001 |
| Group | -0.375487 | 0.1608933 | 53.2 | -0.698167, -0.052807 | 0.0234 |
| Time | -0.017833 | 0.0277749 | 54.0 | -0.073518, 0.0378521 | 0.5236 |
| Group×Time | 0.0830595 | 0.0346414 | 54.0 | 0.0136077, 0.1525112 | 0.0200 |
| Age | 0.0043429 | 0.0071454 | 52.0 | -0.009995, 0.0186812 | 0.5460 |
| Sex | 0.1085662 | 0.1724137 | 52.0 | -0.237407, 0.4545396 | 0.5317 |
| **IL-1β** |  |  |  |  |  |
| Intercept | 1.2618031 | 0.6154942 | 52.0 | 0.0267393, 2.4968669 | 0.0454* |
| Group | -0.15048 | 0.1984832 | 52.4 | -0.548691, 0.2477308 | 0.4518 |
| Time | 0.0078992 | 0.0200263 | 54.0 | -0.032251, 0.0480494 | 0.6948 |
| Group×Time | -0.007298 | 0.0249771 | 54.0 | -0.057374, 0.042778 | 0.7713 |
| Age | -0.009743 | 0.0088488 | 52.0 | -0.027499, 0.0080133 | 0.2759 |
| Sex | -0.325958 | 0.2135146 | 52.0 | -0.754406, 0.1024904 | 0.1329 |

**Table S4.** Associations between inflammatory mediators at baseline for both groups.

|  | **CRP** | **IL-6** | **IL-8** | **IL-10** | **TNF-α** | **IL-1β** |
| --- | --- | --- | --- | --- | --- | --- |
| CRP | — |  |  |  |  |  |
| IL-6 | 0.13 | — |  |  |  |  |
| IL-8 | 0.01 | 0.42 | — |  |  |  |
| IL-10 | -0.05 | 0.68 | 0.32 | — |  |  |
| TNF-α | 0.05 | -0.19 | 0.39 | -0.33 | — | |
| IL-1β | -0.01 | 0.24 | 0.36 | 0.13 | 0.00 | — |

Values are Spearman's correlation coefficients ( $r^2$ ). Values in red denote statistically significant correlations ( $p < 0.05$ )

**Table S5.** Associations between inflammatory mediators pre-post intervention (Δ=concentration T1-concentration T0) for cardiovascular exercise (CE) and standard care (SC) groups.

| **CE** |  |  |  |  |  |  |
| --- | --- | --- | --- | --- | --- | --- |
|  | **CRP** | **IL-6** | **IL-8** | **IL-10** | **TNF-α** | **IL-1β** |
| CRP | — |  |  |  |  |  |
| IL-6 | 0.23 | — |  |  |  |  |
| IL-8 | 0.10 | 0.55 | — |  |  |  |
| IL-10 | -0.03 | 0.59 | 0.24 | — |  |  |
| TNF-α | 0.01 | 0.37 | 0.13 | 0.29 | — | |
| IL-1β | -0.16 | 0.43 | 0.35 | 0.30 | -0.05 | — |
| **SC** |  |  |  |  |  |  |
|  | **CRP** | **IL-6** | **IL-8** | **IL-10** | **TNF-α** | **IL-1β** |
| CRP | — |  |  |  |  |  |
| IL-6 | 0.22 | — |  |  |  |  |
| IL-8 | -0.25 | 0.21 | — |  |  |  |
| IL-10 | 0.05 | 0.49 | 0.53 | – |  |  |
| TNF-α | 0.02 | -0.06 | 0.70 | 0.54 | – | |
| IL-1β | 0.00 | 0.42 | 0.20 | 0.68 | 0.10 | – |

Values are Spearman's correlation coefficients. Values in red denote statistically significant correlations ( $p < 0.05$ )

**Table S6.** Associations between inflammatory mediators pre-post intervention (Δ=concentration T1-concentration T0) and changes in cardio-respiratory fitness (VO_2_ peak), cognition (MoCA), degree of neurological (NIHSS) and motor (Fugl-Meyer) deficit for cardiovascular exercise (CE) and standard care (SC) groups.

| CE |  |  |  |  |  |  |
| --- | --- | --- | --- | --- | --- | --- |
| | CRP | IL-6 | IL-8 | IL-10 | TNF-α | IL-1β |
| $VO_{2peak}$ | -0.03 | -0.35 | -0.16 | -0.09 | -0.01 | 0.90 |
| MoCA | 0.02 | 0.15 | -0.19 | 0.17 | -0.09 | -0.49 |
| NIHSS | -0.19 | 0.11 | -0.30 | 0.17 | -0.01 | 0.34 |
| Fugl-Meyer | 0.15 | -0.52 | -0.16 | -0.57 | 0.36 | 0.15 |
| SC |  |  |  |  |  |  |
| | CRP | IL-6 | IL-8 | IL-10 | TNF-α | IL-1β |
| $VO_{2peak}$ | -0.40 | 0.40 | 0.02 | -0.14 | -0.12 | -0.70 |
| MoCA | 0.10 | -0.24 | -0.42 | -0.35 | -0.17 | -0.82 |
| NIHSS | -0.89 | -0.42 | 0.25 | 0.19 | -0.31 | 0.11 |
| Fugl-Meyer | 0.16 | 0.33 | -0.12 | -0.06 | 0.37 | 0.10 |

Values are Spearman's correlation coefficients. Values in red denote statistically significant correlations ( $p < 0.05$ )

## Supplementary Figures Captions

**Figure S1.**
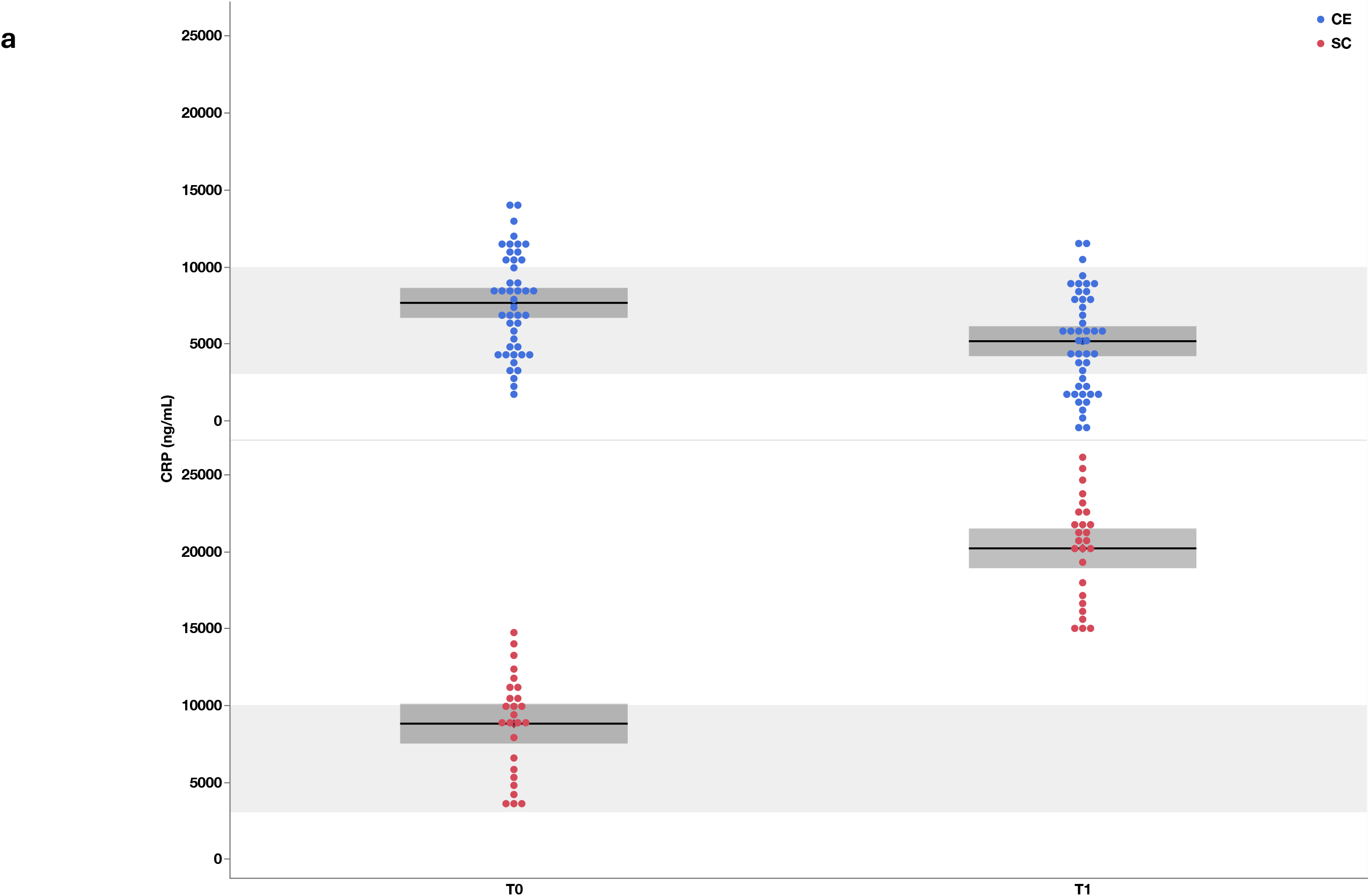
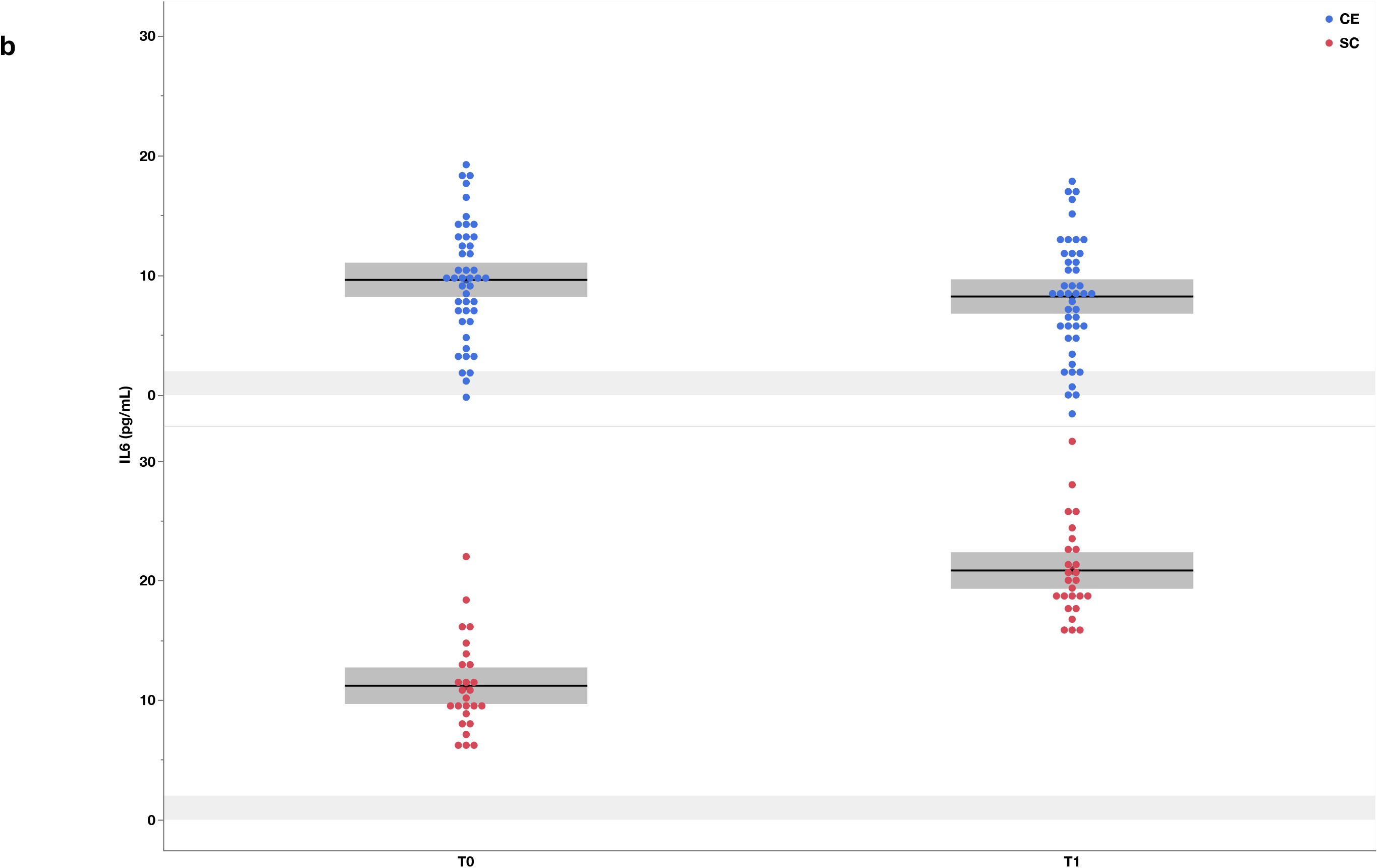
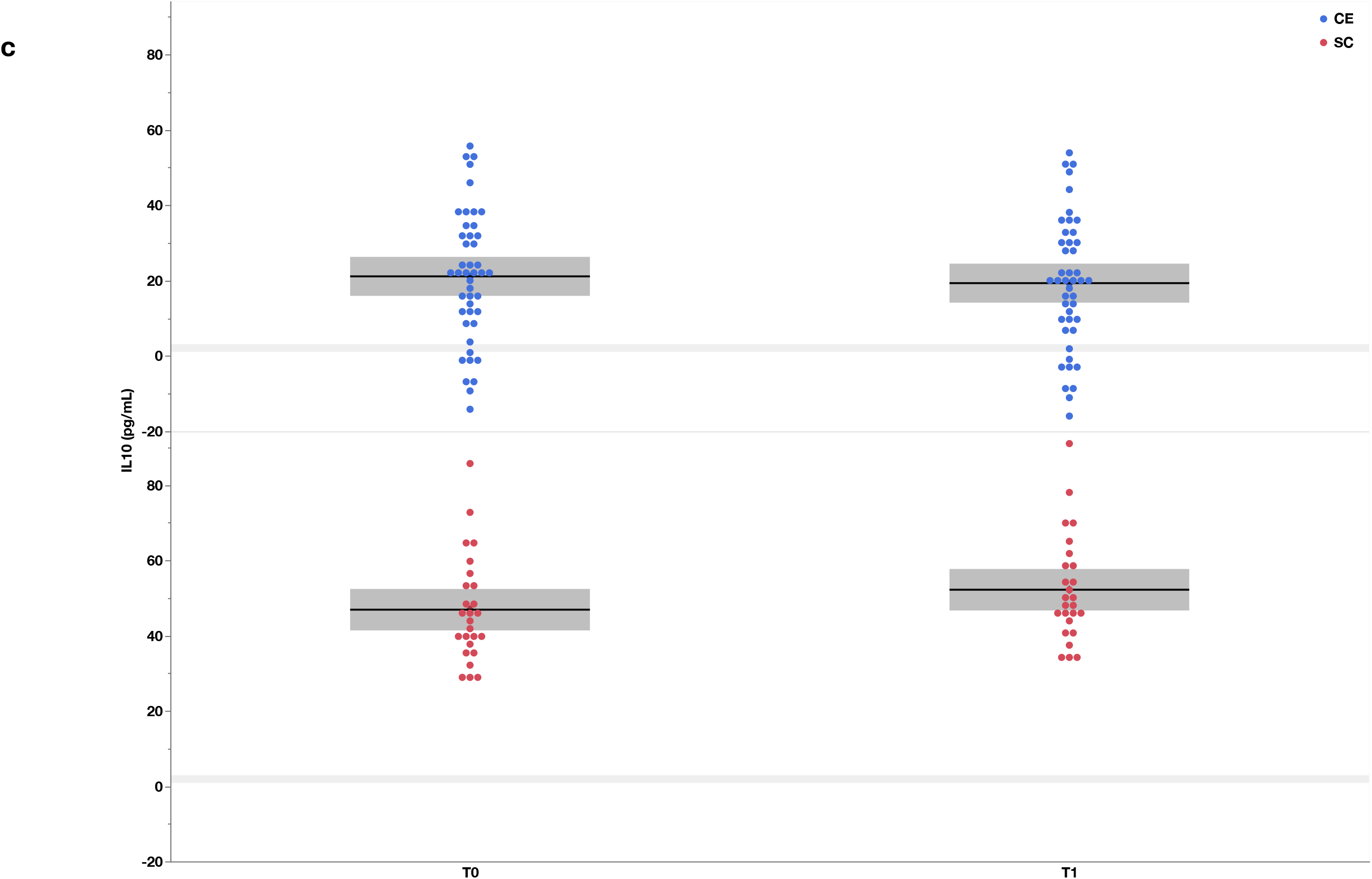
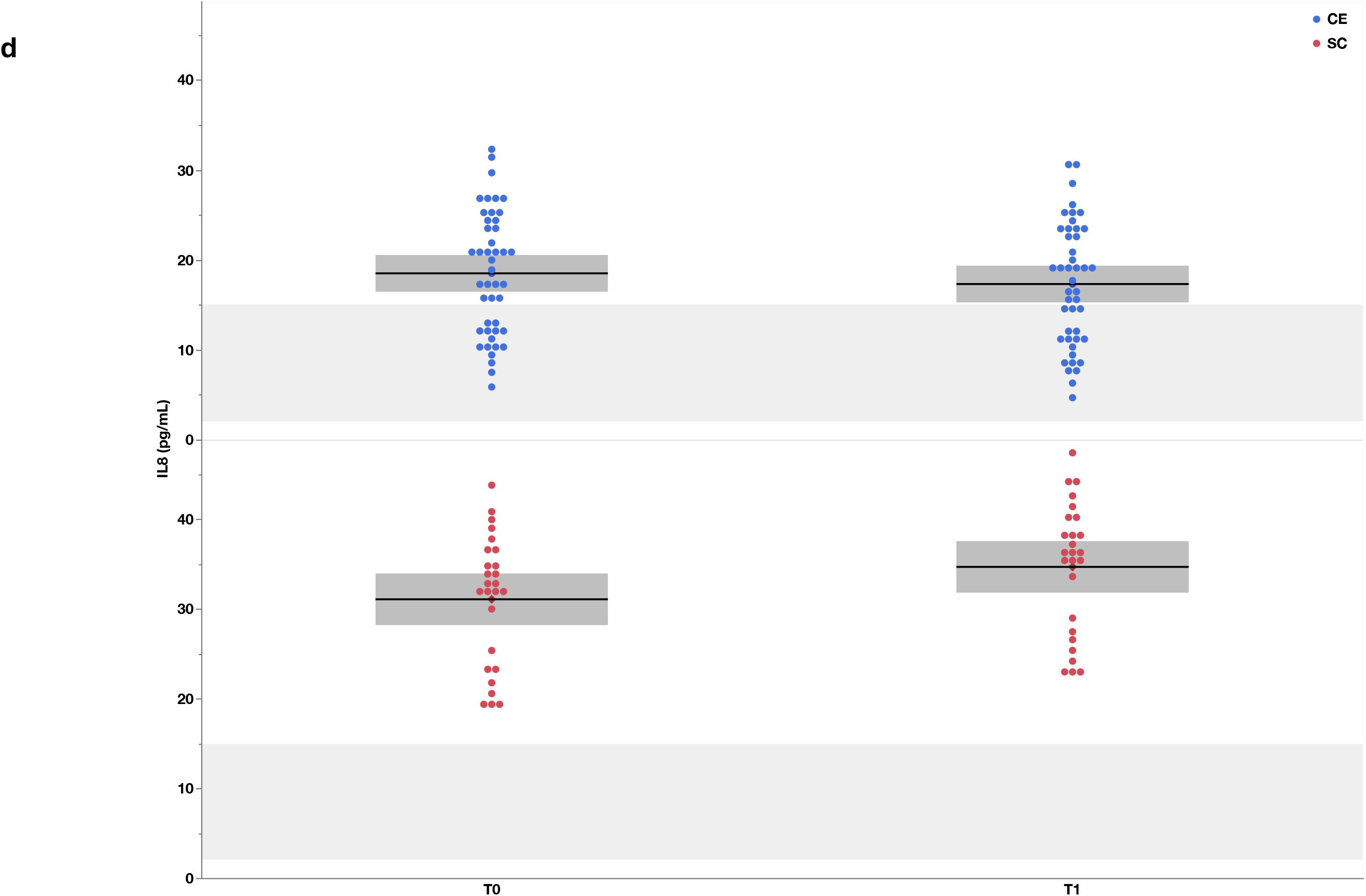
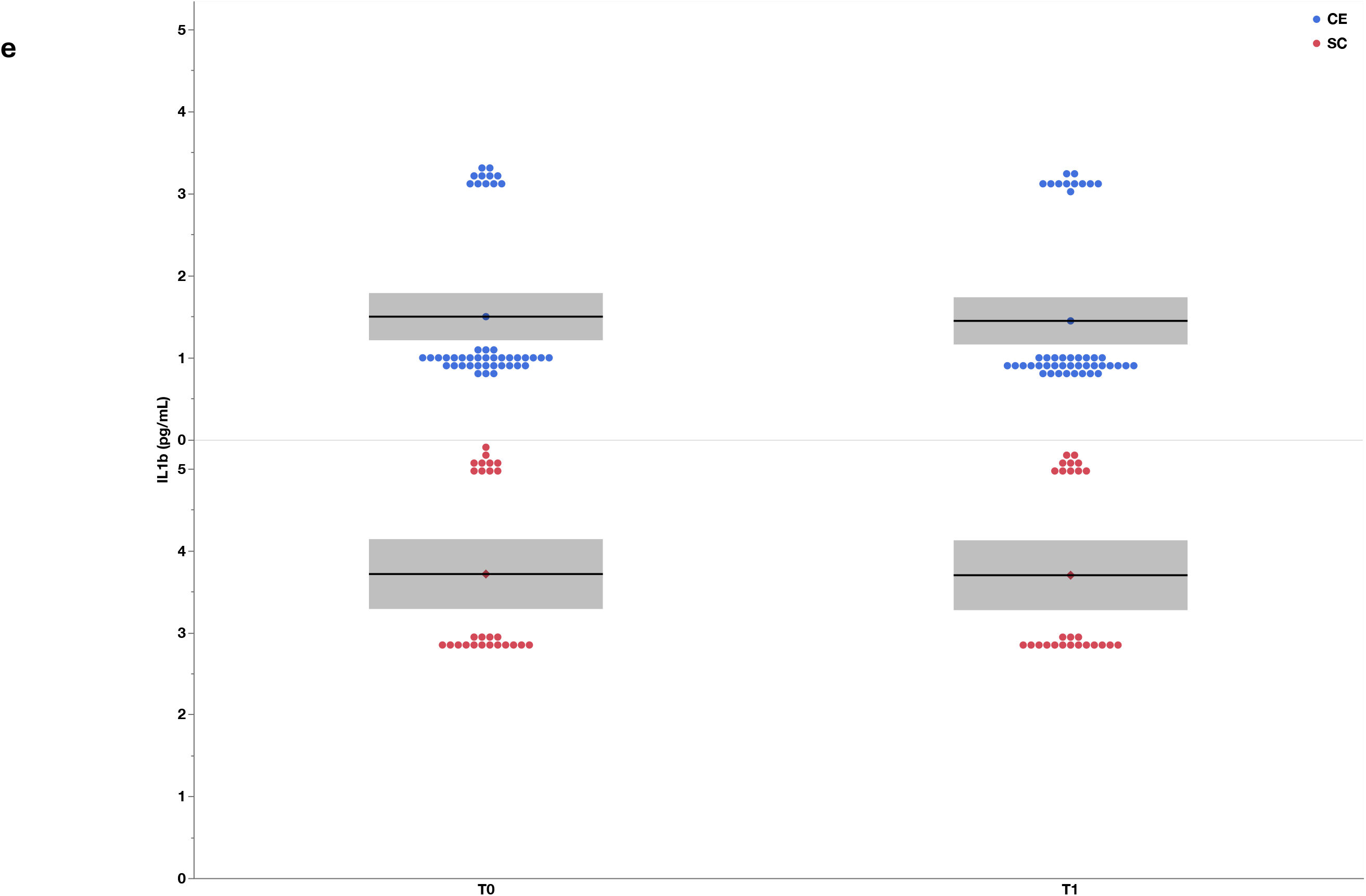
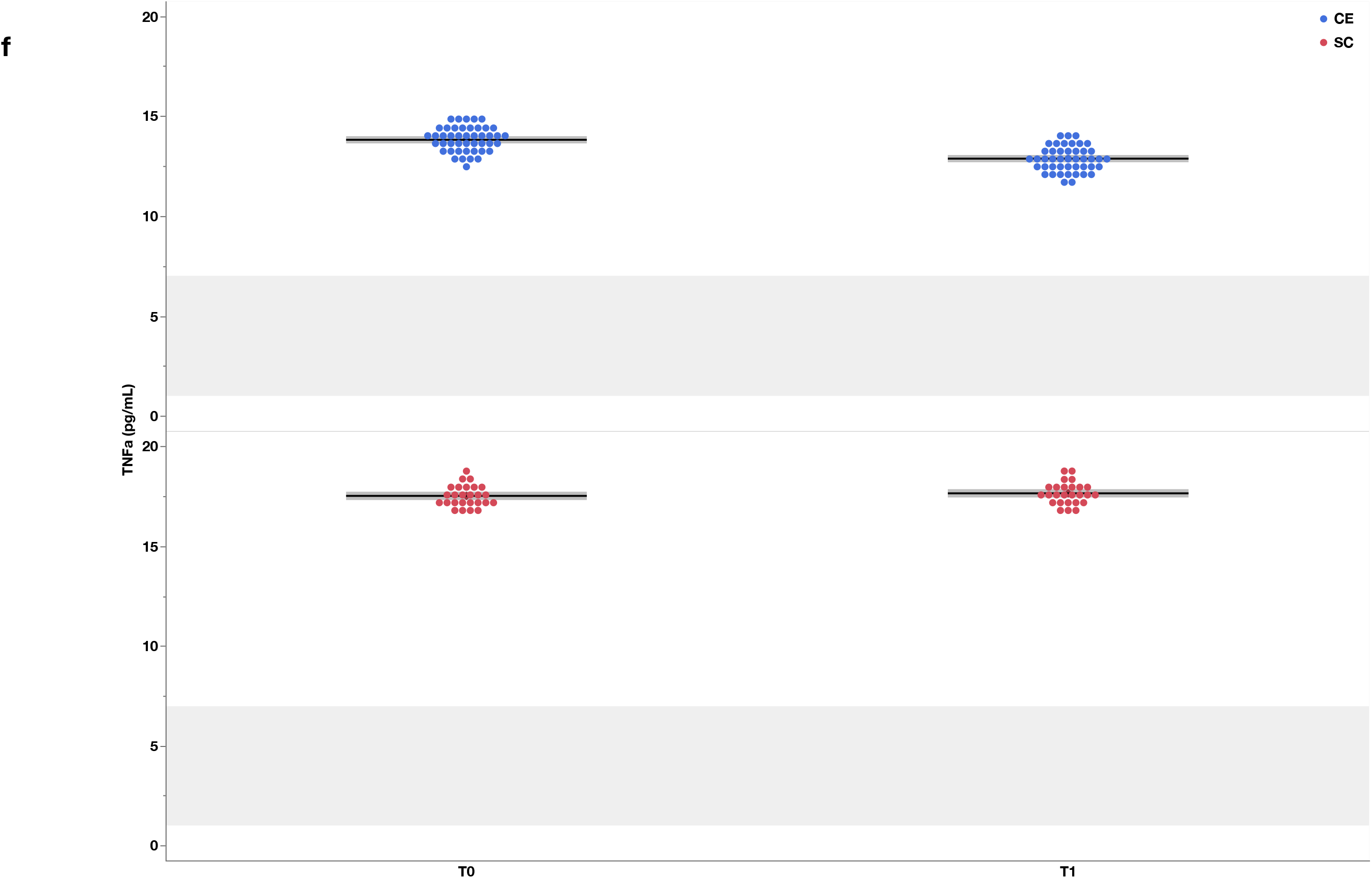
**a-f**. Least square mean estimates plots showing changes in the concentration of the inflammatory mediators CRP, IL-6, IL-8, IL-10, TNF-α and IL-1β for CE and SC from T0 to T1. Values are non-log-transformed predicted mean estimates with 95% CIs. Shaded grey areas represent reference concentration ranges obtained from studies in which the concentration of these mediators was assessed in serum. Ranges for IL-1β are not provided due to the lack of consistent data. These ranges are only approximations and should be interpreted cautiously.

## References

ACSM, 2024. ACSM’s guidelines for exercise testing and prescription. Lippincott Williams & Wilkins, Philadelphia.

Aref, H.M.A., Fahmy, N.A., Khalil, S.H., Ahmed, M.F., ElSadek, A., Abdulghani, M.O., 2020. Role of interleukin-6 in ischemic stroke outcome. The Egyptian Journal of Neurology, Psychiatry and Neurosurgery 56, 12.

Băcilă, C.I., Vlădoiu, M.G., Văleanu, M., Moga, D.F., Pumnea, P.M., 2025. The Role of IL-6 and TNF-Alpha Biomarkers in Predicting Disability Outcomes in Acute Ischemic Stroke Patients. Life (Basel) 15.

Beavers, K.M., Brinkley, T.E., Nicklas, B.J., 2010. Effect of exercise training on chronic inflammation. Clin Chim Acta 411, 785–793.

Bernhardt, J., Hayward, K.S., Kwakkel, G., Ward, N.S., Wolf, S.L., Borschmann, K., Krakauer, J.W., Boyd, L.A., Carmichael, S.T., Corbett, D., Cramer, S.C., 2017. Agreed Definitions and a Shared Vision for New Standards in Stroke Recovery Research: The Stroke Recovery and Rehabilitation Roundtable Taskforce. Neurorehabil Neural Repair 31, 793–799.

Biernaskie, J., Chernenko, G., Corbett, D., 2004. Efficacy of rehabilitative experience declines with time after focal ischemic brain injury. J Neurosci 24, 1245–1254.

Bitencourt, A.C.S., Aveiro, M.E.S., Timoteo, R.P., Bazan, R., Carvalho, E.E.V., Luvizutto, G.J., 2025. Exercise-Induced Modulation of Inflammatory Biomarkers After Stroke: A Systematic Review and Meta-Analysis. J Manipulative Physiol Ther 48, 373–384.

Brough, D., Rothwell, N.J., Allan, S.M., 2015. Interleukin-1 as a pharmacological target in acute brain injury. Exp Physiol 100, 1488–1494.

Calabrese, F., Rossetti, A.C., Racagni, G., Gass, P., Riva, M.A., Molteni, R., 2014. Brain-derived neurotrophic factor: a bridge between inflammation and neuroplasticity. Front Cell Neurosci 8, 430.

Carmichael, N.S., Deijnen, H.R., Wong, S.Y., Williams, T.O., Kontopantelis, E., Cowie, L., Jones, E., Drag, L., Buckwalter, M.S., Grainger, J.R., Allan, S.M., Smith, C.J., 2026. Longitudinal plasma interleukin-6 and post-stroke cognitive outcomes: The Stroke-IMPaCT study. Alzheimers Dement 22, e71261.

Charlson ME, P.P., Ales KL, MacKenzie CR., 1987. A new method of classifying prognostic comorbidity in longitudinal studies: development and validation. J Chronic Dis. 40, 373–383.

Clausen, B.H., Wirenfeldt, M., Hogedal, S.S., Frich, L.H., Nielsen, H.H., Schroder, H.D., Ostergaard, K., Finsen, B., Kristensen, B.W., Lambertsen, K.L., 2020. Characterization of the TNF and IL-1 systems in human brain and blood after ischemic stroke. Acta Neuropathol Commun 8, 81.

Couch, C., Mallah, K., Borucki, D.M., Bonilha, H.S., Tomlinson, S., 2022. State of the science in inflammation and stroke recovery: A systematic review. Ann Phys Rehabil Med 65, 101546.

da Cunha, M.J., Pires Dorneles, G., Peres, A., Maurer, S., Horn, K., Souza Pagnussat, A., 2024. tDCS does not add effect to foot drop stimulator and gait training in improving clinical parameters and neuroplasticity biomarkers in chronic post-stroke: randomized controlled trial. Int J Neurosci 134, 1518–1527.

De Las Heras, B., Rodrigues, L., Cristini, J., Moncion, K., Ploughman, M., Tang, A., Fung, J., Roig, M., 2024. Measuring Neuroplasticity in Response to Cardiovascular Exercise in People With Stroke: A Critical Perspective. Neurorehabil Neural Repair 38, 303–321.

De Las Heras, B., Rodrigues, L., Cristini, J., Weiss, M., Prats-Puig, A., Roig, M., 2022. Does the Brain-Derived Neurotrophic Factor Val66Met Polymorphism Modulate the Effects of Physical Activity and Exercise on Cognition? Neuroscientist 28, 69–86.

De Las Heras, B., Rodrigues, L., Cristini, J., Yu, E., Gan-Or, Z., Arbour, N., Thiel, A., Tang, A., Fung, J., Eng, J.J., Roig, M., 2025. Investigating the Acute and Chronic Effects of Cardiovascular Exercise on Brain-Derived Neurotrophic Factor in Early Subacute Stroke. Neurorehabil Neural Repair 39, 653–665.

Di Filippo, M., Sarchielli, P., Picconi, B., Calabresi, P., 2008. Neuroinflammation and synaptic plasticity: theoretical basis for a novel, immune-centred, therapeutic approach to neurological disorders. Trends Pharmacol Sci 29, 402–412.

Dong, Y., Sharma, V.K., Chan, B.P., Venketasubramanian, N., Teoh, H.L., Seet, R.C., Tanicala, S., Chan, Y.H., Chen, C., 2010. The Montreal Cognitive Assessment (MoCA) is superior to the Mini-Mental State Examination (MMSE) for the detection of vascular cognitive impairment after acute stroke. J. Neurol. Sci. 299, 15–18.

Dromerick, A.W., Geed, S., Barth, J., Brady, K., Giannetti, M.L., Mitchell, A., Edwardson, M.A., Tan, M.T., Zhou, Y., Newport, E.L., Edwards, D.F., 2021. Critical Period After Stroke Study (CPASS): A phase II clinical trial testing an optimal time for motor recovery after stroke in humans. Proc Natl Acad Sci U S A 118.

Geng, H.H., Wang, X.W., Fu, R.L., Jing, M.J., Huang, L.L., Zhang, Q., Wang, X.X., Wang, P.X., 2016. The Relationship between C-Reactive Protein Level and Discharge Outcome in Patients with Acute Ischemic Stroke. Int J Environ Res Public Health 13.

Gertz, K., Kronenberg, G., Kälin, R.E., Baldinger, T., Werner, C., Balkaya, M., Eom, G.D., Hellmann-Regen, J., Kröber, J., Miller, K.R., Lindauer, U., Laufs, U., Dirnagl, U., Heppner, F.L., Endres, M., 2012. Essential role of interleukin-6 in post-stroke angiogenesis. Brain 135, 1964–1980.

Grayston, A., Baptista, M., Wemyss, K., Taylor, R., Cullen, G., Jafree, S.S., Luka, N., Cox, J.R., Konkel, J.E., Brough, D., Allan, S.M., Pinteaux, E., 2025. Specific deletion of interleukin-1 beta in microglia improves acute outcome and modulates neurogenesis after ischemic stroke. bioRxiv, 2025.2012.2019.695391.

Hsu, C.-C., Tsai, H.-H., Fu, T.-C., Wang, J.-S., 2019. Exercise Training Enhances Platelet Mitochondrial Bioenergetics in Stroke Patients: A Randomized Controlled Trial. Journal of Clinical Medicine 8.

Iordache, M.P., Buliman, A., Costea-Firan, C., Gligore, T.C.I., Cazacu, I.S., Stoian, M., Teoibaș-Şerban, D., Blendea, C.D., Protosevici, M.G., Tanase, C., Popa, M.L., 2025. Immunological and Inflammatory Biomarkers in the Prognosis, Prevention, and Treatment of Ischemic Stroke: A Review of a Decade of Advancement. Int J Mol Sci 26.

Kelly, P.J., Lemmens, R., Tsivgoulis, G., 2021. Inflammation and Stroke Risk: A New Target for Prevention. Stroke 52, 2697–2706.

Kirzinger, B., Stroux, A., Rackoll, T., Endres, M., Floel, A., Ebinger, M., Nave, A.H., 2021. Elevated Serum Inflammatory Markers in Subacute Stroke Are Associated With Clinical Outcome but Not Modified by Aerobic Fitness Training: Results of the Randomized Controlled PHYS-STROKE Trial. Front Neurol 12, 713018.

Kleiner, G., Marcuzzi, A., Zanin, V., Monasta, L., Zauli, G., 2013. Cytokine levels in the serum of healthy subjects. Mediators Inflamm 2013, 434010.

Krawcyk, R.S., Vinther, A., Petersen, N.C., Faber, J., Iversen, H.K., Christensen, T., Lambertsen, K.L., Rehman, S., Klausen, T.W., Rostrup, E., Kruuse, C., 2019. Effect of Home-Based High-Intensity Interval Training in Patients With Lacunar Stroke: A Randomized Controlled Trial. Front Neurol 10, 664.

Kriz, J., Lalancette-Hébert, M., 2009. Inflammation, plasticity and real-time imaging after cerebral ischemia. Acta Neuropathologica 117, 497–509.

Kwah, L.K., Diong, J., 2014. National Institutes of Health Stroke Scale (NIHSS). J. Physiother. 60, 61.

Lai, Y.J., Hanneman, S.K., Casarez, R.L., Wang, J., McCullough, L.D., 2019. Blood biomarkers for physical recovery in ischemic stroke: a systematic review. Am J Transl Res 11, 4603–4613.

Lin JH, H.I., Sheu CF, Hsieh CL., 2004. Psychometric properties of the sensory scale of the Fugl-Meyer Assessment in stroke patients. Clinical Rehabilitation 18, 391–397.

Martinez de Toda, I., Gonzalez-Sanchez, M., Diaz-Del Cerro, E., Valera, G., Carracedo, J., Guerra-Perez, N., 2023. Sex differences in markers of oxidation and inflammation. Implications for ageing. Mech Ageing Dev 211, 111797.

McCabe, J.J., Walsh, C., Gorey, S., Arnold, M., DeMarchis, G.M., Harris, K., Hervella, P., Iglesias-Rey, R., Jern, C., Katan, M., Li, L., Miyamoto, N., Montaner, J., Purroy, F., Rothwell, P.M., Stanne, T.M., Sudlow, C., Ueno, Y., Vicente-Pascual, M., Whiteley, W., Woodward, M., Kelly, P.J., 2024. Interleukin-6, C-Reactive Protein, and Recurrence After Stroke: A Time-Course Analysis of Individual-Participant Data. Stroke 55, 2825–2834.

Moncion, K., Rodrigues, L., De Las Heras, B., Wiley, E., Sikorska, K., Cristini, J., Allison, E.Y., Eng, J., Tang, A., Roig, M., 2025. A Whole-Body Exercise Test to Assess Cardiorespiratory Fitness Across the Stroke Recovery Continuum. Med Sci Sports Exerc.

Muller, S., Kufner, A., Dell’Orco, A., Rackoll, T., Mekle, R., Piper, S.K., Fiebach, J.B., Villringer, K., Floel, A., Endres, M., Ebinger, M., Nave, A.H., 2021. Evolution of Blood-Brain Barrier Permeability in Subacute Ischemic Stroke and Associations With Serum Biomarkers and Functional Outcome. Front Neurol 12, 730923.

Murphy, T.H., Corbett, D., 2009. Plasticity during stroke recovery: from synapse to behaviour. Nat Rev Neurosci 10, 861–872.

Oliveira, D.M.G., Aguiar, L.T., de Oliveira Limones, M.V., Gomes, A.G., da Silva, L.C., de Morais Faria, C.D.C., Scalzo, P.L., 2019. Aerobic Training Efficacy in Inflammation, Neurotrophins, and Function in Chronic Stroke Persons: A Randomized Controlled Trial Protocol. J Stroke Cerebrovasc Dis 28, 418–424.

Pacinella, G., Bona, M.M., Todaro, F., Ciaccio, A.M., Daidone, M., Tuttolomondo, A., 2025. Tracing Inflammation in Ischemic Stroke: Biomarkers and Clinical Insight. Int J Mol Sci 26.

Pandey, A., Patel, M.R., Willis, B., Gao, A., Leonard, D., Das, S.R., Defina, L., Berry, J.D., 2016. Association Between Midlife Cardiorespiratory Fitness and Risk of Stroke: The Cooper Center Longitudinal Study. Stroke 47, 1720–1726.

Petryshen, T.L., Sabeti, P.C., Aldinger, K.A., Fry, B., Fan, J.B., Schaffner, S.F., Waggoner, S.G., Tahl, A.R., Sklar, P., 2010. Population genetic study of the brain-derived neurotrophic factor (BDNF) gene. Mol Psychiatry 15, 810–815.

Piepke, M., Clausen, B.H., Ludewig, P., Vienhues, J.H., Bedke, T., Javidi, E., Rissiek, B., Jank, L., Brockmann, L., Sandrock, I., Degenhardt, K., Jander, A., Roth, V., Schadlich, I.S., Prinz, I., Flavell, R.A., Kobayashi, Y., Renne, T., Gerloff, C., Huber, S., Magnus, T., Gelderblom, M., 2021. Interleukin-10 improves stroke outcome by controlling the detrimental Interleukin-17A response. J Neuroinflammation 18, 265.

Poorhabibi, H., Weiss, K., Rosemann, T., Knechtle, B., Eslami, R., Tartibian, B., Tayebi, S.M., Sheikhhoseini, R., 2025. Short-Lived Exercise-Induced Exerkines Modulate Inflammation for Chronic Disease Prevention: A Systematic Review and Meta-Analysis. Biomolecules 15.

Sandvig, H.V., Aam, S., Alme, K.N., Askim, T., Beyer, M.K., Ellekjaer, H., Ihle-Hansen, H., Lydersen, S., Mollnes, T.E., Munthe-Kaas, R., Naess, H., Saltvedt, I., Seljeseth, Y.M., Thingstad, P., Wethal, T., Knapskog, A.B., 2023. Plasma Inflammatory Biomarkers Are Associated With Poststroke Cognitive Impairment: The Nor-COAST Study. Stroke 54, 1303–1311.

Schmidt-Pogoda, A., Ruck, T., Strecker, J.K., Hoppen, M., Fazio, L., Vinnenberg, L., Maus, B., Wachsmuth, L., Cerina, M., Diederich, K., Lichtenberg, S., Abberger, H., Haertel, L., Schafflick, D., Meyer Zu Horste, G., Herrmann, A.M., Hundehege, P., Narayanan, V., Nelke, C., Kruithoff, K., Bosbach, J., Vicari, E., Ramcke, T., Beuker, C., Hadaschik, E., Budde, T., Faber, C., Wiendl, H., Hansen, W., Meuth, S.G., Minnerup, J., 2025. Exercise facilitates post-stroke recovery through mitigation of neuronal hyperexcitability via interleukin-10 signaling. Nat Commun 16, 8928.

Serra, M.C., Hafer-Macko, C.E., Robbins, R., O’Connor, J.C., Ryan, A.S., 2022. Randomization to Treadmill Training Improves Physical and Metabolic Health in Association With Declines in Oxidative Stress in Stroke. Arch Phys Med Rehabil 103, 2077–2084.

Shaheen, H.A., Daker, L.I., Abbass, M.M., Abd El Fattah, A.A., 2018. The relationship between the severity of disability and serum IL-8 in acute ischemic stroke patients. Egypt J Neurol Psychiatr Neurosurg 54, 26.

Shi, K., Tian, D.C., Li, Z.G., Ducruet, A.F., Lawton, M.T., Shi, F.D., 2019. Global brain inflammation in stroke. Lancet Neurol 18, 1058–1066.

Sobowale, O.A., Parry-Jones, A.R., Smith, C.J., Tyrrell, P.J., Rothwell, N.J., Allan, S.M., 2016. Interleukin-1 in Stroke: From Bench to Bedside. Stroke 47, 2160–2167.

Stahmeyer, J.T., Stubenrauch, S., Geyer, S., Weissenborn, K., Eberhard, S., 2019. The Frequency and Timing of Recurrent Stroke: An Analysis of Routine Health Insurance Data. Dtsch Arztebl Int 116, 711–717.

Sun, W., Wang, S., Nan, S., 2021. The Prognostic Determinant of Interleukin-10 in Patients with Acute Ischemic Stroke: An Analysis from the Perspective of Disease Management. Dis Markers 2021, 6423244.

Tayebi, S.M., Poorhabibi, H., Heidary, D., Amini, M.A., Sadeghi, A., 2025. Impact of aerobic exercise on chronic inflammation in older adults: a systematic review and meta-analysis. BMC Sports Sci Med Rehabil 17, 229.

Washburn, R.A., Zhu, W., McAuley, E., Frogley, M., Figoni, S.F., 2002. The physical activity scale for individuals with physical disabilities: Development and evaluation. Arch. Phys. Med. Rehabil. 83, 193–200.

Winbeck, K., Poppert, H., Etgen, T., Conrad, B., Sander, D., 2002. Prognostic relevance of early serial C-reactive protein measurements after first ischemic stroke. Stroke 33, 2459–2464.

Xiao, L., Huang, Y., Wu, L., Zeng, S., Qiu, C., Li, X., Xie, L., Wu, D., 2025. The role of inflammation in Ischemic stroke: from biomarker to treatment. Frontiers in Immunology Volume 16 - 2025.

Zhou, X., Fragala, M.S., McElhaney, J.E., Kuchel, G.A., 2010. Conceptual and methodological issues relevant to cytokine and inflammatory marker measurements in clinical research. Curr Opin Clin Nutr Metab Care 13, 541–547.

Zhu, H., Hu, S., Li, Y., Sun, Y., Xiong, X., Hu, X., Chen, J., Qiu, S., 2022. Interleukins and Ischemic Stroke. Front Immunol 13, 828447.

Zietz, A., Gorey, S., Kelly, P.J., Katan, M., McCabe, J.J., 2024. Targeting inflammation to reduce recurrent stroke. Int J Stroke 19, 379–387.

